# Video-based Detection of Delirium in Hospitalized Adults

**DOI:** 10.64898/2026.05.11.26352902

**Authors:** Maanasa Mendu, Ryan A. Tesh, Kyle Pellerin, Grace E. Steward, Ivo H. Cerda, Marta Williams, Mia Colman, Simran Shah, Alice D. Lam, Sydney S. Cash, M. Brandon Westover, Eyal Y. Kimchi

## Abstract

Delirium, a dynamic neuropsychiatric condition associated with morbidity and mortality, remains underdiagnosed due to reliance on subjective, intermittent screening tools. Objective and potentially continuous identification is needed to improve clinical care. We developed and validated an analytic framework for delirium classification based on automatically extracted video features. In this prospective cohort study, patients (≥ 18 years) admitted to the inpatient medical or neurological ward of a tertiary academic center between August 2020 and March 2022 with an expected stay longer than one night were enrolled. Daily structured delirium assessments and brief video recordings were performed in consenting patients. Videos were analyzed using deep learning pose estimation to extract keypoints and calculate behavioral features based on eye, face, and limb postures and movements. Four machine learning models (logistic regression, gradient boosting, support vector machines, and random forests) were trained to predict delirium status from extracted features. Model performance was evaluated on 20 repetitions of three-fold cross-validation using the area under the curve of the receiver operating characteristics curve (AUC ROC). The cohort included 109 videos from 25 male and 25 female participants (median age: 72, IQR: 63.25-78). Twenty videos (18%) were from patients with delirium. Keypoints for this dataset were more accurately extracted using a customized ResNet-101 model developed with DeepLabCut (sensitivity 0.94, specificity 0.89, compared to human-labeled gold standards) than using off-the-shelf models. Keypoints were then used to generate behavioral features summarizing movement and postures throughout the video. A support vector machine model achieved an average delirium classification AUC ROC of 0.79 (SD ± 0.09), sensitivity of 0.71 (SD ± 0.16), and specificity of 0.78 (SD ± 0.07). This study demonstrates the feasibility of identifying delirium using brief videos in clinically heterogeneous cohorts and reveals novel features for objective identification.

**Author Summary:** Delirium is a sudden change in attention and awareness that commonly affects hospitalized patients. It is linked with longer hospital stays, cognitive decline, and death. Patients with delirium often show changes in movements and behaviors such as slowed movement, restlessness, or excessive scanning of the environment. Since current screening tools rely on intermittent human interactions, they can be subjective and miss the fluctuating nature of delirium, leading to underdiagnosis. We sought to explore whether short video recordings could be used to detect delirium automatically. In our study, we enrolled 50 hospitalized patients and conducted daily delirium assessments and video recordings. We used a machine learning model to analyze patients’ eye movements, facial expressions, and body postures. We found that video-derived features could be used to identify delirium in a small clinical cohort. While needing further validation in outside cohorts, this study shows an important proof-of-concept for objective delirium monitoring in heterogeneous clinical contexts without adding burden to clinical staff.

## Introduction

Delirium is a neuropsychiatric condition characterized by acute and fluctuating changes in attention and awareness [1] that affects more than 20% of hospitalized patients over 65 years old [2]. Delirium has significant associations with increased mortality, long-term cognitive decline [3], and higher healthcare costs [4]. Therefore, timely detection of this acute and dynamic condition is important.

Despite high prevalence, delirium remains underdiagnosed [5]. An ideal biomarker would be easy to measure in vulnerable populations, rooted in pathophysiology, and able to capture fluctuations in status over time. While the gold standard for delirium diagnosis is clinical assessment according to expert-consensus criteria [1], this is limited by the availability of expert clinicians [6]. Over 30 delirium assessment tools have therefore been developed as alternatives to clinical interview [7,8], but while these tools can be used by clinical staff or caregivers to identify delirium, they involve subjective interpretation of intermittent interactions.

Electroencephalography (EEG) is considered a promising tool for monitoring delirium, but its use can be limited by available expertise in application and interpretation [6]. Additionally, standard clinical EEG systems have the potential to impact patient experience negatively over long periods of monitoring due to skin irritation, restricted movement, and increased anxiety [9–11], which could exacerbate a patient’s risk for delirium.

Automated video-based monitoring of vulnerable patients for behaviors indicative of delirium could address limitations of EEG while maintaining advantages of continuous, objective monitoring. Patients with delirium are known to exhibit changes in psychomotor activity, such as hypoactivity and slowed movement, or hyperactivity, including restlessness, picking and grasping, and excessive environment scanning [12]. Subjective observations of these motor behaviors are already key features of existing delirium assessments and trainings [13–15]. While previous studies have investigated actigraphy [16], electro-oculograms [17], and motor function assessments as potential indicators of delirium [18], they measure a limited scope of behavior and still involve placing devices on patients.

Recent advancements in deep learning have enabled the automated recognition of patient motor behaviors, patient emotions, and whether a patient is supervised by hospital staff [19–21]. One previous study demonstrated the feasibility of using video data to monitor delirium status, albeit in a small intensive care unit (ICU) cohort of fewer than 20 patients [22], while a similar study is ongoing [23]. Additionally, it has been shown that pose estimation of neonatal ICU patients could successfully predict EEG diagnoses for sedation and cerebral dysfunction [24]. These prior studies support our hypothesis that features relevant to delirium can be identified from video. In this study, we aimed to determine whether machine learning analysis of brief patient videos could identify delirium in a larger, more clinically heterogenous adult population outside of the ICU.

## Methods

This manuscript follows the Transparent Reporting of a Multivariable Prediction Model for Individual Prognosis or Diagnosis+AI (TRIPOD+AI) expanded checklist guidelines [25] (included in Supplementary Material).

### Ethics Statement

This study was approved by the Institutional Review Board of Massachusetts General Hospital and the Partners Human Research Committee (#2020P000678). Formal written consent was obtained from patients or Legally Authorized Representatives when patients did not have capacity to consent.

### Study Design and Participants

We conducted a single center, prospective observational cohort study to develop and validate predictive biomarkers of delirium (ClinicalTrials.gov, Identifier: NCT04602988). To capture the heterogeneity of delirium, all patients admitted to inpatient medical and neurological units at Massachusetts General Hospital were approached for potential inclusion, depending on staff availability. The broader overall study occurred from August 2020 to March 2022. Starting on November 13, 2020, participants were asked whether they would provide additional, optional consent for video recording of daily delirium evaluations. The results reported here are based on the subset of patients who personally or through Legally Authorized Representatives consented to video recordings. Eligible participants were 18 years or older and had an expected length of stay of at least one night. Patients were excluded if they had head wounds that would interfere with EEG placement, due to requirements for the concurrent parent study. Non-English-speaking patients were excluded due to incomplete translation of the employed delirium assessments. To enhance clinical inclusiveness otherwise, no other exclusion criteria were used, allowing enrollment of patients with neurologic illness, cognitive impairment, and alcohol or substance use disorders. Patients were evaluated daily up to 7 days while inpatient, pending staff availability. Demographic data was collected from the electronic medical record.

### Delirium Evaluation

Patients were assessed for delirium using the 3-Minute Diagnostic Interview for Confusion Assessment Method (3D-CAM) [26]. Delirium evaluations typically occurred in the afternoon to minimize disruption of ongoing clinical care. All research staff administering assessments underwent rigorous training via didactics, video cases, supervised assessments, and ongoing quality monitoring for consistency.

### Video Data Collection

Videos were recorded during patient delirium evaluations using a GoPro HERO7 Black with a frame rate of 30 frames per second at a resolution of 1920×1440 pixels. The camera was mounted at the foot of the hospital bed using two adjustable neck extensions (total ∼40 cm) to elevate it to patient height, focused on the patient.

### Video Data Validation

To ensure collected video data was appropriate for pose estimation, all videos were screened for quality assurance. Patients in the Epilepsy Monitoring Unit were excluded due to head wraps that extended over the cheeks to under the jaw, as were fully masked patients due to occlusion of facial keypoints. Additionally, videos were excluded due to any data errors, such as file corruption, truncation, or blank frames.

### Overall Machine Learning Framework

We used a multiple-stage computational framework (Supplementary Figure 1). After video recordings, we identified keypoints for eyes, face, torso, and limbs in every frame, using human pose estimation algorithms. After frame-by-frame keypoint identification, behavioral features were constructed across entire videos. Summary values of behavioral features for each video were then used to train, test, and cross-validate delirium prediction models.

#### Human Pose Estimation Model Development for Keypoint Identification

A total of 3 human pose estimation approaches were assessed. We selected two publicly available, pretrained models, developed and validated externally, from Google’s MediaPipe suite: one with high density facial representation (FaceMesh), and one with representation of limbs (BlazePose) [27].

We developed the third model based on ResNet-101, a 101 layer network pre-trained on ImageNet, using DeepLabCut (Supplementary Methods) [28]. Approximately 10 frames were selected via k-means clustering from each patient video. For each frame, 35 keypoints (22 facial and 13 extremity) were labeled by team members using a guide to ensure consistency (Supplementary Table 1). The most experienced rater reviewed all labeled keypoints to ensure consistency.

An initial DeepLabCut model was then trained starting with a restricted version of the dataset, which included 314 frames from 35 patients. Of these data, 80% of frames were used to train the model, and 20% of frames from these initial patients were held out for internal testing (Supplementary Table 2). Generalizability to novel videos was then assessed on a held-out data set of 86 frames from 10 videos not previously included in the data set. Based on incomplete generalization performance (see Results), a second, larger, comprehensive model was developed with a total of 782 frames from 109 videos of all 50 patients, using the same internal 80:20 train-test-ratio.

#### Pose Estimation Model Evaluation

The accuracies of the pose estimation models were assessed using Euclidean distance, sensitivity, and specificity of identified keypoints compared to ground truth human labels. Pose estimation algorithms BlazePose and DeepLabCut predict a visibility confidence for point identification. Presence vs absence was determined by applying thresholds to each keypoint. Keypoint thresholds were chosen by optimizing the F2 score for each model and body region on training data only. This threshold was then applied to testing data to determine sensitivity (percentage of human-labeled keypoints identified by each model) and specificity (percentage of keypoints correctly omitted when absent/not human-labeled, e.g., if not visible due to being turned away or occluded). The best-performing keypoint estimation model was selected for keypoint inference for all full-length patient videos.

#### Behavioral Feature Extraction

Following keypoint inference, we calculated 58 behavioral features in the domains of eye, head, mouth, and extremity movements selected through a combination of qualitative literature review and visual inspection of videos (Supplementary Table 3). Example eye movement features included pupil movements, including both horizontal and vertical, and changes in the horizontal and vertical eyelid axes. Blinks were scored according to the rates over time, durations, percent of frames with eyes open, and the relative vertical eyelid gap. Head features were calculated as changes in orientation such as roll, pitch, yaw, and nose orientation. Mouth features included movements of each of the lips and percentage of frames with the mouth open. Extremity features included arm movements and relative positions between hands, wrist, elbows, and shoulders, and movements of the knees (see Supplementary Table 4 for definitions). Each behavioral feature was then summarized across all the frames of the video, using the mean, median, standard deviation, and median absolute deviation (4 summary measures per feature), yielding 232 features per video.

#### Delirium Classification Model Training and Testing

Due to the complex, non-linear relationships between extracted features and delirium status, we employed machine learning classification to predict delirium. Conventional machine learning was selected over deep learning because research shows the former performs better with limited sample sizes and tabular feature data [29]. Key types of supervised machine learning models: XGBoost (XGB) [30], random forest (RF), logistic regression (LR), and support vector machine (SVM), were chosen for their performance on binary classification, interpretability, and their ability to handle class imbalance.

To identify the best performing classifier, we used repeated k-fold cross-validation, performing 20 repetitions of three-fold cross-validation. This approach ensured that each fold contained enough positive delirium cases in both training and validation sets, thereby improving the reliability and generalizability of our performance estimates in the absence of a held-out test set. Each repetition of cross-validation used different random seeds to split the data, allowing us to evaluate how sensitive model performance was to changes in the data.

Each cross-validation set was generated using StratifiedGroupKFold from scikit-learn 1.8.0, which maintains similar class ratios across folds, while additionally ensuring that data from participants with multiple samples is kept strictly in only one of either the training or test set. Within each fold, we first applied one of several feature selection methods (minimum Redundancy Maximum Relevance or mRMR [31], Lasso regression selection, Mutual Information, chi-square statistic, or none), then trained the model on the selected features, and only afterwards applied both feature selection and model estimation to fold test data. Model performance was calculated as the mean area under the curve of the receiver operating characteristic curves (AUC ROC) across folds and repetitions for each algorithm-feature⍰selection combination, with variability reported using the standard deviation.

Due to the limited number of samples available, we did not tune hyperparameters within the cross-validation folds to avoid overfitting. Instead, we selected hyperparameters tuned to binary classification that would balance the weight of the minority class and protect from overfitting, such as limiting the maximum depth of tree-based classifiers (Supplementary Table 5). All other hyperparameters were kept at their default settings.

Performance metrics are reported as the mean across all cross-validation folds and repetitions (20 repetitions with three folds each, yielding a mean of 60 values). The standard deviation across those same values is reported to quantify algorithm sensitivity to training on different subsets of the dataset.

To test whether the selected models out-performed models based on shuffled data, we conducted a permutation analysis using 500 permutations [32]. For each permutation, the delirium status labels were randomly sampled without replacement on the full dataset. Each permuted dataset was then processed using the pipelines described above to obtain a distribution of AUC ROC scores. We compared the resulting distribution of scores for each algorithm-feature⍰selection combination to the observed score from the non-permuted data. The p-value was calculated as the proportion of permuted scores that exceeded the performance of the model trained with the true class labels.

#### Explainability

In order to understand which behavioral features drove model predictions, we calculated Shapley additive explanations, a model-agnostic metric that quantifies associations between features and model output, via the associated toolbox [33]. For each cross-validation fold, SHAP values were computed for all test observations. Feature-level SHAP values were then averaged across all applicable observations within each algorithm and feature-selection method. Observations where a feature was not selected for the fitted model were assigned a NaN value to be excluded from the average. The top 10 features were those with the highest absolute mean SHAP values across iterations within each algorithm and feature-selection method combination.

### Statistical Analyses

Numerical data are reported as medians with interquartile ranges unless otherwise stated. Categorical variables are reported as counts or frequencies. Groups were compared via the Kruskal Wallis test and chi-squared tests. To compare pose-estimation models while accounting for clustered repeated measures, we used a hierarchical linear mixed-effects model (lme4 in R) of log-transformed pixel errors, given their non-normal distributions prior to transformation [34]. Random effects were nested such that pixel-error was attributed to image frame embedded within videos from patients. Model appropriateness was evaluated by assessing residuals and heteroscedasticity. Given the clustered nature of the data, a sandwich estimator of the variance was used to calculate 95% confidence intervals (CI) for sensitivity and specificity, using a logistic regression model with clustering by image [35]. All test statistics with a p-value less than 0.05 were deemed statistically significant, with corrections for multiple comparisons, except where indicated for descriptive data. A complete list of tools and packages used in model development and statistical analysis is at https://github.com/KimchiLab/samba_delirium_videos_release.

## Results

### Participant Characteristics

The analyzed dataset consisted of 109 recordings from 50 patients (Figure 1), with most participants contributing one or two videos (n=35 of 50, 70%, Supplementary Figure 2). The point prevalence of delirium diagnosis was 18.5% (20/109) of video evaluations. These videos were selected from a larger dataset of consenting patients after stepwise exclusion of patients in the Epilepsy Monitoring Unit (40 videos excluded from 11 patients), then exclusion of videos from patients wearing masks (40 videos excluded from 21 patients), and lastly exclusion of videos with file errors (3 videos excluded from 3 patients, Figure 1). Analysis of descriptive cohort data revealed no significant differences in age, primary service, length of stay, or disposition of patients with or without delirium (Table 1, p-values were not corrected for multiple comparisons given the descriptive nature of this data). Among the videos included, 20 videos were performed at times of positive delirium assessments from 17 patients. Videos had a median duration of 3.2 [2.5-4.9] minutes, consistent with the expected 3 minute duration of 3D-CAM assessments [26].

**Figure 1.**
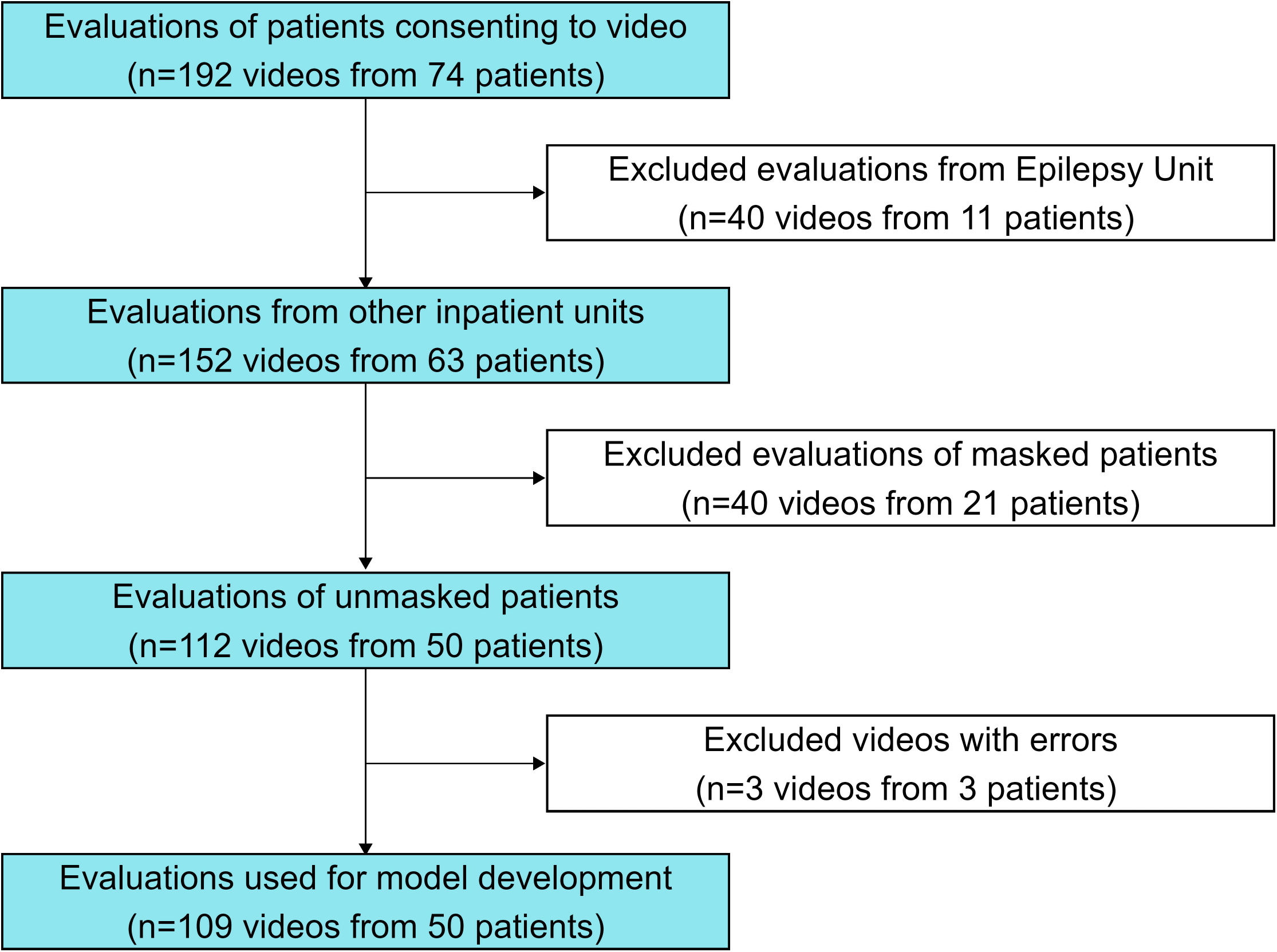
Flowchart of Video Evaluations and Patient Cohort. The study video dataset included videos of delirium evaluations from consenting patients. In stepwise fashion, we first excluded all patients recorded in the Epilepsy Monitoring Unit (40 videos excluded from 11 patients). From the remaining videos of patients in other units, we then excluded videos from patients when they were wearing masks (40 videos excluded from 21 patients). Lastly, a small number of videos were excluded due to video file errors (3 videos excluded from 3 patients). The number of videos excluded at each stage is labeled on the right-hand side of the figure. Because patients could contribute more than one video, exclusion of a video at one stage did not necessarily mean exclusion of that patient entirely.

**Table 1.** Patient cohort characteristics.

|  | Overall | Never Delirious | Delirious at Least Once | p |
| --- | --- | --- | --- | --- |
| Number of Patients | 50 | 33 | 17 |  |
| Total Video Evaluations (number videos during positive delirium evaluations) | 109 (20) | 73 (0) | 36 (20) |  |
| Number of video evaluations per patient | 2.0 [1.0, 3.0] | 2.0 [1.0, 3.0] | 2.0 [1.0, 3.0] | 0.715 |
| Age (years) | 72.00<br>[63.25, 78.00] | 70.00<br>[60.00, 76.00] | 74.00<br>[68.00, 81.00] | 0.073 |
| Gender (Female) | 25 (50.0%) | 17 (51.5%) | 8 (47.1%) | 1.000 |
| Race and Ethnicity |  |  |  | 0.396 |
| Hispanic or Latino | 1 (2.0%) | 1 (3.0%) | 0 (0.0%) |  |
| Native Hawaiian or Other Pacific Islander | 1 (2.0%) | 0 (0.0%) | 1 (5.9%) |  |
| White | 47 (94.0%) | 31 (93.9%) | 16 (94.1%) |  |
| Other | 1 (2.0%) | 1 (3.0%) | 0 (0.0%) |  |
| Hypoactive Patients (RASS < 0) | 6 (12.0%) | 1 (3.0%) | 5 (29.4%) | 0.024 |
| Length of Stay (days) | 6.0 [4.0, 8.0] | 5.0 [3.0, 7.0] | 7.0 [4.0, 9.0] | 0.122 |
| Primary Clinical Service |  |  |  |  |
| Medicine | 39 (78.0%) | 26 (78.8%) | 13 (76.5%) | 1.000 |
| Neurology | 11 (22.0%) | 7 (21.2%) | 4 (23.5%) |  |
| Disposition |  |  |  |  |
| Death | 2 (4.0%) | 1 (3.0%) | 1 (5.9%) | 0.212 |
| Home | 37 (74.0%) | 27 (81.8%) | 10 (58.8%) |  |
| Rehabilitation | 11 (22.0%) | 5 (15.2%) | 6 (35.3%) |  |

Patient characteristics are shown for the overall cohort, then for patients never diagnosed with delirium during their study assessments, followed by those diagnosed with delirium at least once during study assessments. Delirium was defined using the 3-Minute Diagnostic Interview for Confusion Assessment Method (3D-CAM). Numerical variables are reported as medians with interquartile range (IQR) presented in brackets, and statistical significance is calculated using the rank-sum test. Categorical variables are reported as counts with percentages in parentheses, and statistical significance is calculated using the Chi-squared test. P-values were not corrected for multiple comparisons given the descriptive nature of this data.

### Human Pose Estimation Model Performance

An initial DeepLabCut model developed with a restricted dataset of 314 frames achieved a median error of 1.39 pixels and 2.25 pixels for training and test data respectively and significantly outperformed FaceMesh and BlazePose models (Figure 2A). However, when evaluated on a held-out validation set of 86 frames from 10 unseen patient videos, the restricted model’s performance was statistically similar to FaceMesh for facial points, and more variable for extremity points but still statistically different compared to BlazePose (Figure 2A).

**Figure 2.**
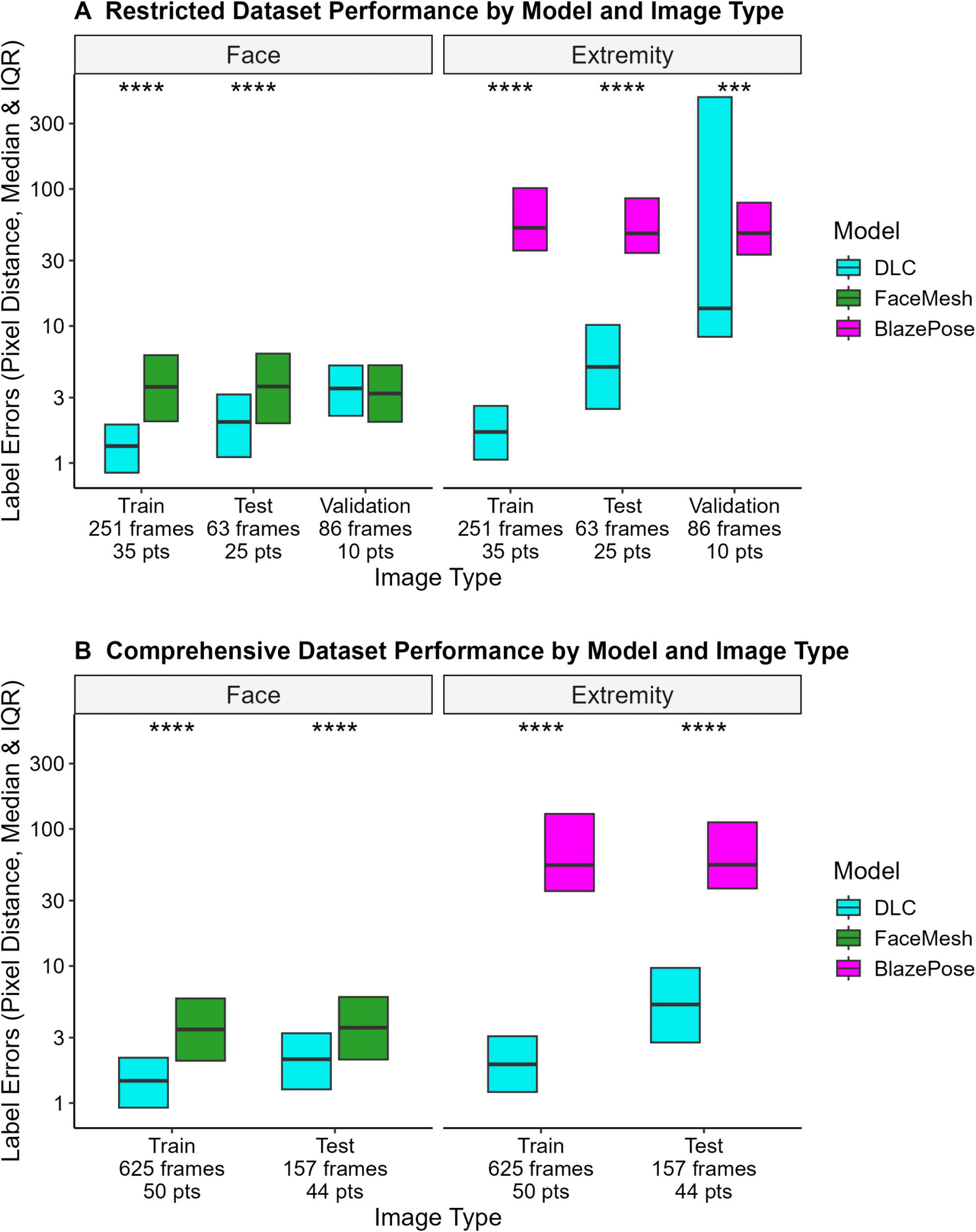
Identification of patient keypoints by a trained computer vision models, compared to general purpose models. We compared keypoint estimation for three different computer vision models: a DeepLabCut network trained on labeled frames and two general purpose models (FaceMesh and BlazePose). Frame and patient counts (pts) are representative of totals in each dataset. A. The boxplots display the model errors (on a log scale) for an initial restricted dataset (n = 400 frames, 45 unique patients). The restricted dataset was divided into training, testing, and validation sets for training, internally evaluating, and assessing the generalizability of the DLC model. The same videos could be included in both train and test portions of the dataset, but a unique set of videos was used for validation. Error was calculated using the Euclidean distance formula between the model-predicted points and ground truth labels. Lower values mean better keypoint accuracy. Box elements correspond to center line=median, lower limit=25th percentile, upper limit=75th percentile. Statistical significance was calculated using a linear mixed effects model with log transformed distance values, with fixed effects of Model type, Data portion, and Body region. Nested random effects included patients, video recordings, and selected frames. Prespecified post-hoc comparisons were calculated using Sidak correction for multiple comparisons. The DLC model outperformed FaceMesh for training and testing datasets (**** p < 0.0001) but was similar for validation data (p > 0.05). The DLC model significantly outperformed BlazePose on all datasets (**** p < 0.0001, *** p < 0.001). B. The boxplots display the model errors (on a log scale) on the comprehensive dataset (n = 782 frames, 50 unique patients). For this model, frames from all patients were used to train and test the model, therefore only training and test portions are available for analysis. The DLC model had significantly lower error for all comparisons of the Comprehensive Dataset, significantly outperforming both BlazePose and FaceMesh models on both training and testing frames (**** p < 0.0001). Conventions are as in A.

Therefore, we optimized the DeepLabCut model with an expanded dataset of 782 frames from 50 patients. This comprehensive model’s performance remained significantly better than both FaceMesh and BlazePose for facial and extremity points respectively (p < 0.0001, Figure 2B). Furthermore, the comprehensive model had the highest overall sensitivity (0.94, 95% CI 0.92-0.96, from 3748/3983 keypoints) and specificity (0.89, 95% CI 0.86-0.91, from 924/1041 keypoints) for identification of non-training keypoints, including both facial and extremity keypoints (Supplementary Table 6). Further characterization of the comprehensive model’s performance showed similar performance by body region across gender (Supplementary Table 7) and delirium status (Supplementary Table 8). Due to its performance, this model was used to identify keypoints from all frames of all videos and then generate behavioral features for each video for further analysis.

### Differences in Individual Features Between Patients With and Without Delirium

Out of 232 calculated behavioral video features, 23 were found to be significantly different between patients with or without delirium (p < 0.05, without correction for multiple comparisons given the descriptive nature of this presentation) (Supplementary Table 9). A notable theme from these features was that patients with delirium often had irregular movements of the pupil, eyes, mouth, and extremities. These individual features did not remain significant after correction for multiple comparisons but provided context for the following multivariable, cross-validated classification.

### Video-Feature Based Delirium Classification

The machine learning pipeline with the best performance classifying delirium was Support Vector Machine classification using all features, yielding an average AUC ROC of 0.79 (SD ± 0.09), sensitivity of 0.71 (SD ± 0.16), specificity of 0.78 (SD ± 0.07), positive predictive value of 0.42 (SD ± 0.09), and negative predictive value of 0.92 (SD ± 0.04) (Figure 3A, Supplementary Table 10). The other three algorithms’ (Logistic Regression, XGBoost, and Random Forest) top feature selection models all performed with a mean AUC ROC score of 0.69 or greater (Figure 3B-D). Statistical significance was confirmed against a null distribution of randomly shuffled class permutations (p < 0.002, Supplementary Figure 3). Support Vector Machine classification qualitatively outperformed other machine learning algorithms trained and tested on the same feature sets (Supplementary Table 10).

**Figure 3.**
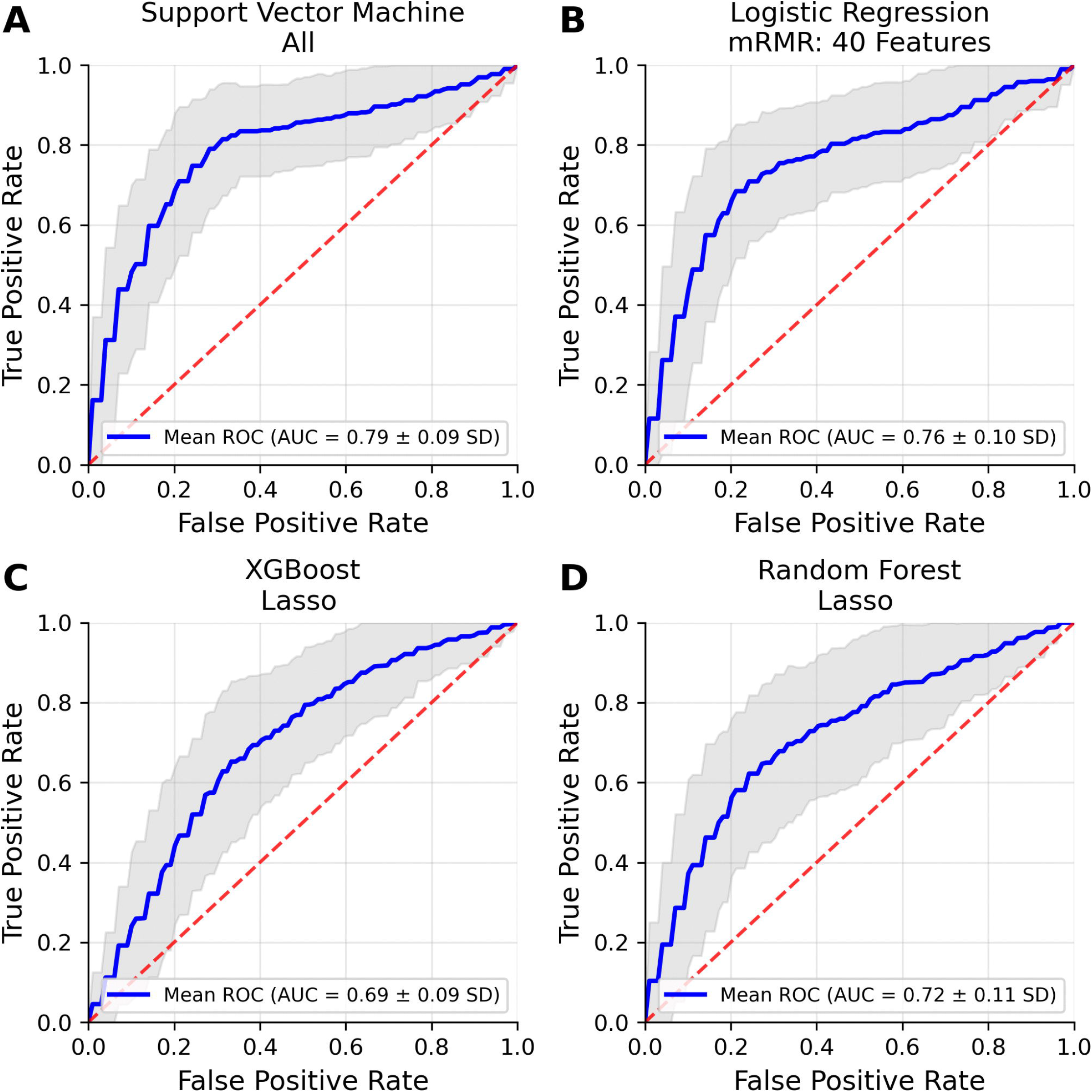
Delirium Prediction Using Machine Learning Analysis of Video-Based Features. The receiver operating characteristic (ROC) curves of the best-performing algorithm-feature-selection combinations show efficacy for the use of video-based features for the objective classification of delirium. A. The Support Vector Machine algorithm fitted with all features performed the best out of all algorithm-feature-combinations and had a mean area under the curve (AUC) of 0.79 (SD ±0.09). Logistic Regression (B), Extreme Gradient Boosting (C), and Random Forest (D) algorithms all performed with a mean AUC ROC at or above 0.69. The shaded grey area is representative of the standard deviation (SD) based on 20 repetitions of three-fold cross-validation, while the dashed red line illustrates the performance of a classifier based on random chance.

We further investigated the features used by the best performing model using Shapley analysis. Features related to variability in elbow movements and blink duration were most prominent in the top 10 features (Figure 4). The model also made use of eye opening, knee movements, and wrist movements, highlighting the variety of information used to classify delirium. An analysis of the other top performing model combinations showed repeated importance of extremity movement and standard deviation of blink duration (Supplementary Figure 4).

**Figure 4.**
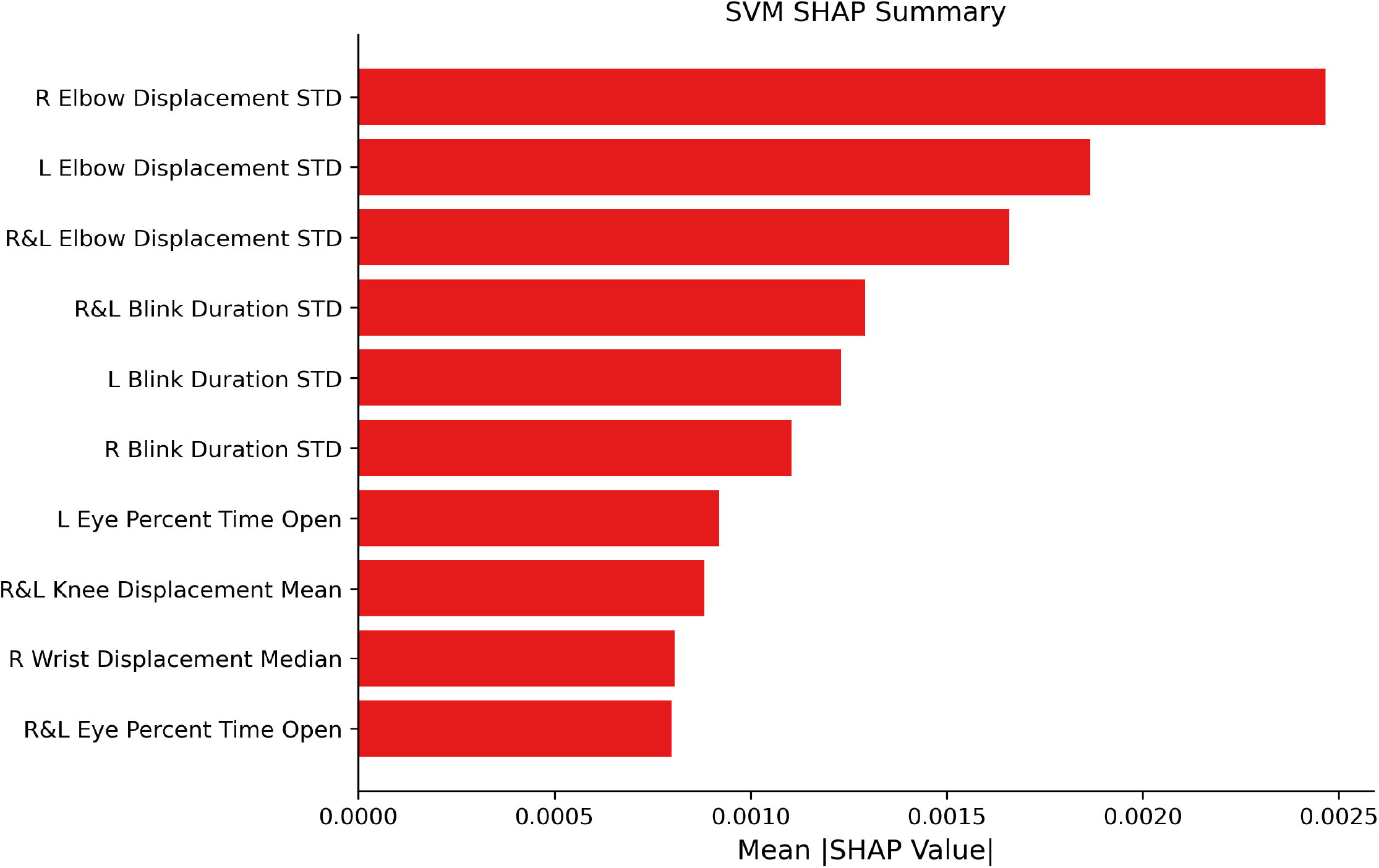
Shapley Explanatory Analysis of Selected Features. The top 10 features used in the Support Vector Machine algorithm (SVM) ranked in descending order of importance by their mean absolute Shapley analysis (SHAP) values across folds and repetitions. Features of the highest importance clustered around the upper extremities, such as the standard deviation of elbow displacement (variability in elbow movements), and standard deviation of blink duration (variability in blink duration). Displacement was measured as the frame-by-frame change in position for the noted keypoints, with summary measures being taken across all changes between frames. “R” denotes right, “L” denotes left, and “R&L” denotes that the metric was averaged across both sides to calculate this feature. “STD” denotes standard deviation. See Supplementary Table 4 for further descriptions of how each feature was calculated, and Supplementary Figure 4 for Shapley analysis results from additional algorithms.

## Discussion

We developed a machine learning model to classify delirium status using automatically extracted keypoints and features from brief videos of hospitalized patients. Our findings indicate that non-invasive video-based analysis has the potential to classify delirium status with approximately three minutes of video data.

We first evaluated the feasibility of keypoint estimation in videos of hospitalized adults. Off-the-shelf keypoint estimation models trained on human fitness pose data, such as BlazePose and FaceMesh, did not match the performance of a customized model, particularly with regard to limbs. Variations in lighting and bed position, as well as obstacles common in the hospital environment (e.g., trays, bedding, and medical equipment) are absent from the training datasets of these general-purpose models. Prior research demonstrates the importance of domain specific adaptation of human pose estimation to clinical settings such as fine-tuning patient-specific CNNs and applying temporal filtering [36,37]. This suggests that off-the-shelf models are unlikely to be as accurate or capable of producing video features clinically relevant to the evaluation of hospitalized patients, especially in the context of delirium where motor activity is particularly relevant [38].

Consistent with these findings, fine-tuning a custom model to perform keypoint estimation on patient videos using DeepLabCut, independent of delirium status, improved performance. While our original model trained on a restricted dataset did not generalize well to out-of-sample videos, a second iteration trained on a larger, comprehensive dataset was more accurate. This suggests that hospital-specific or patient-specific pose models are needed to produce clinically relevant video features. We cannot currently determine the generalizability of this model generated to test hypotheses for subsequent delirium classification in this dataset, but we suspect that larger, more diverse datasets of patient videos would be required to make a more robust and generalizable hospital environment specific pose estimation model. Future work should focus on the collection of such datasets, ideally in a collaborative framework and implementing newer pose models with temporal filtering [39]. But because keypoint estimation was developed independent of delirium status, with similar accuracy for patients with and without delirium, we proceeded with this model to assess whether accurate keypoint estimation can help classify delirium.

We considered several combinations of different machine learning algorithms and feature selection methods to classify delirium. We found that a support vector machine algorithm fitted with all features had the highest test performance. This combination was advantageous due to attributes of the support vector machine algorithm, which is especially useful for analyzing high-dimensional data and working with small to medium datasets, due to its robustness to both noise and overfitting [40].

Analysis of individual features combined with the feature selection processes demonstrated the importance of upper extremity and eye related features in delirium prediction. Although prior studies have not quantitatively profiled specific limb movements in patients with delirium, research has demonstrated an association between delirium and motor function [41]. Wrist actigraphy in post-operative patients showed lower mean activity one day post-op in patients with delirium compared to those without delirium [42], and nighttime actigraphy data (dynamic time warping) was a significant predictive factor for post-stroke delirium in patients with intracerebral hemorrhage [43]. A pilot study of continuous video and accelerometry data in the surgical ICU found that patients with delirium exhibited greater wrist and lower-extremity movement but lower upper extremity movements across a 24-hour period [44]. In the context of structured clinical interviews, eye movement and upper extremity movements may have been reflective of inattention. Additionally, inappropriate placement of limbs may be indicative of reduced environmental awareness. There likely is a nuanced relationship between motor function and delirium status as prior studies demonstrate decreased mobility in patients with delirium [45]. Further study may identify more specific associations between behavioral features and subtypes of delirium [46].

Significant behavioral features in delirium can serve as guides for identifying brain regions that may be implicated in its pathophysiology, especially given recent multi-modal studies of neural correlates of unstructured behavior in epilepsy monitoring units [47–49]. Disturbances of oculomotor function and limb movements may reflect disruptions in basal ganglia-thalamocortical circuits responsible for key roles in attention and motor control [50]. The associations between dopamine, Parkinson’s disease, and delirium [51] have suggested a role for the basal ganglia in delirium, but this structure may be understudied using bedside, superficially focused methods such as EEG [52] and near infrared spectroscopy [53]. Different techniques are warranted for continued investigation of these structures, including neuroimaging [54,55] and preclinical models [56].

### Limitations

Our study has several limitations. Firstly, the small cohort of patients from a single center may affect generalizability. The pose-estimation model was trained on all patients, which may overstate generalizability to unseen patients. Other limitations include the camera standardization used for video capture and the class imbalance in the fraction of patients with RASS scores of less than zero. Additionally, video recordings were filmed during clinical delirium assessments, prompting future research as to whether such behavioral features can be applied to other contexts. Another significant limitation was the lack of racial and ethnic diversity in our cohort. This absence of diversity can lead to biased pose estimation and delirium classification models, reducing their applicability to the broader population. Future research should include a more diverse patient population to enhance the generalizability and utility of delirium detection models.

### Implications for Practice and Future Directions

This study demonstrates the feasibility of identifying delirium from brief bedside videos in a heterogeneous cohort. Clinically meaningful tools will require several further developments. Video capture in the hospital can be accomplished several ways: via cameras installed in patient rooms being used for fall detection [57], via dedicated carts [58] or portable cameras such as video-EEG machines [59], or via personal devices such as smartphones; all of which could function as local edge computing devices. Each option carries legal and ethical implications, not only for patients, but also for healthcare providers and surrounding individuals. Procedures and guidelines will be necessary to ensure patient privacy, data minimization, encryption, de⍰identification, and consent. While deep learning directly on videos may be useful with large datasets, storing video data to refine prediction algorithms increases privacy concerns. Sharing keypoint estimates rather than images may mitigate some of the privacy concerns [60]; however, real-time keypoint estimation will require better algorithms that can generalize across diverse clinical settings and cohorts. Such an algorithm could potentially be developed across multiple sites using federated learning.

Previous studies have shown that delirium risk can be predicted by increasingly powerful machine learning algorithms on a variety of data modalities, including video [22,24], EEG [61], and electronic health records [62]. However, training models on single modality datasets has significant disadvantages when compared to the potential development of a multi-modal algorithm. For example, patients with delirium often exhibit inappropriate behavior, such as pulling at dressings and IV lines. While these behaviors may be captured by pose estimation, video-based pose estimation alone may have more difficulty distinguishing between unstructured videos, for example a patient who is asleep at night or one who has hypoactive delirium. While our study shows the promise of video pose estimation during daytime interaction, combination with even limited EEG, electronic health records, and time data could enhance the efficacy of continuous delirium monitoring, while also decreasing reliance on any one data source.

The potential research benefits of video monitoring for delirium would be to serve as digital biomarkers that support more precise phenotyping and facilitate phenotype targeted interventional studies. The potential clinical benefits would include earlier detection of delirium onset, especially for the more prognostically concerning but frequently underdiagnosed hypoactive phenotype [63,64]. An early warning system potentially supported by central monitoring, could flag providers about worsening delirium status. However, current delirium trials are split into prevention trials before the occurrence of delirium, where there is good evidence for the benefit of multimodal nonpharmacologic interventions [65], or treatment trials after the onset of clinically overt delirium, where the evidence for intervention benefit is less clear [66]. Deployment of video monitoring for delirium will ultimately be dependent not only on developing infrastructure, but also on proving that there is a clear evidence-based benefit to intervention based on early video-augmented detection of delirium status.

## Conclusion

Behavioral features extracted from brief patient videos were able to identify delirium in a cohort of hospitalized adults. These findings demonstrate the potential of autonomous delirium classification in vulnerable patients. Future studies should evaluate whether video can predict delirium status continuously, and must validate findings on larger, more diverse cohorts.

## Supporting information

TRIPODAI Checklist

## Data Availability

Anonymized data and supporting documentation for the results reported in this study are available at https://github.com/KimchiLab/samba_delirium_videos_release. This repository stores anonymized demographic data, extracted video features, and individual keypoint data at the individual patient level. For reasons of patient privacy, original video data cannot be made available. From the available data, external users will be able to reproduce the customized features as calculated from DeepLabCut keypoints as well as retrain and revalidate the delirium classification models.

https://github.com/KimchiLab/samba_delirium_videos_release4

## Acknowledgements

The authors would like to acknowledge the patients and their families who contributed to this study, and Dr. Margaret M. Banker, PhD, and the Northwestern University Biostatistics Collaboration Center for biostatistical consultation.

## Data Access, Responsibility, and Analysis

EYK had full access to all the data in the study and takes responsibility for the integrity of the data and the accuracy of the data analysis. MM, GES, & EYK conducted and are responsible for the data analysis.

## Data Sharing Statement

Anonymized data and supporting documentation for the results reported in this study are available at https://github.com/KimchiLab/samba_delirium_videos_release4. This repository stores anonymized demographic data, extracted video features, and individual keypoint data at the individual patient level. For reasons of patient privacy, original video data cannot be made available. From the available data, external users will be able to reproduce the customized features as calculated from DeepLabCut keypoints as well as retrain and revalidate the delirium classification models.

## References

1. American Psychiatric Association. Diagnostic and Statistical Manual of Mental Disorders, 5th Edition: DSM-5. 5 edition. Washington, D.C: American Psychiatric Publishing; 2013.

2. Bellelli G, Morandi A, Di Santo SG, Mazzone A, Cherubini A, Mossello E, et al. “Delirium Day”: a nationwide point prevalence study of delirium in older hospitalized patients using an easy standardized diagnostic tool. BMC Med. 2016;14: 106. doi:10.1186/s12916-016-0649-8

3. Davis DHJ, Muniz Terrera G, Keage H, Rahkonen T, Oinas M, Matthews FE, et al. Delirium is a strong risk factor for dementia in the oldest-old: a population-based cohort study. Brain. 2012;135: 2809–2816. doi:10.1093/brain/aws190

4. Leslie DL. One-Year Health Care Costs Associated With Delirium in the Elderly Population. Arch Intern Med. 2008;168: 27. doi:10.1001/archinternmed.2007.4

5. Geriatric Medicine Research Collaborative. Delirium is prevalent in older hospital inpatients and associated with adverse outcomes: results of a prospective multi-centre study on World Delirium Awareness Day. BMC Med. 2019;17: 229. doi:10.1186/s12916-019-1458-7

6. Wiegand TLT, Rémi J, Dimitriadis K. Electroencephalography in delirium assessment: a scoping review. BMC Neurol. 2022;22: 86. doi:10.1186/s12883-022-02557-w

7. De J, Wand APF. Delirium Screening: A Systematic Review of Delirium Screening Tools in Hospitalized Patients. GERONT. 2015;55: 1079–1099. doi:10.1093/geront/gnv100

8. Helfand BKI, D’Aquila ML, Tabloski P, Erickson K, Yue J, Fong TG, et al. Detecting Delirium: A Systematic Review of Identification Instruments for NON-ICU Settings. J American Geriatrics Society. 2021;69: 547–555. doi:10.1111/jgs.16879

9. Oliveira AS, Schlink BR, Hairston WD, König P, Ferris DP. Proposing Metrics for Benchmarking Novel EEG Technologies Towards Real-World Measurements. Front Hum Neurosci. 2016;10. doi:10.3389/fnhum.2016.00188

10. Casson AJ. Wearable EEG and beyond. Biomed Eng Lett. 2019;9: 53–71. doi:10.1007/s13534-018-00093-6

11. Marathe M. Inpatient Experiences of Long-Term Monitoring: The Case of EEG. Companion Publication of the 2024 Conference on Computer-Supported Cooperative Work and Social Computing. New York, NY, USA: Association for Computing Machinery; 2024. pp. 511–518. doi:10.1145/3678884.3681899

12. Morandi A, Di Santo SG, Cherubini A, Mossello E, Meagher D, Mazzone A, et al. Clinical Features Associated with Delirium Motor Subtypes in Older Inpatients: Results of a Multicenter Study. The American Journal of Geriatric Psychiatry. 2017;25: 1064–1071. doi:10.1016/j.jagp.2017.05.003

13. Trzepacz PT, Mittal D, Torres R, Kanary K, Norton J, Jimerson N. Validation of the Delirium Rating Scale-revised-98: comparison with the delirium rating scale and the cognitive test for delirium. J Neuropsychiatry Clin Neurosci. 2001;13: 229–242. doi:10.1176/jnp.13.2.229

14. Meagher D, Adamis D, Leonard M, Trzepacz P, Grover S, Jabbar F, et al. Development of an abbreviated version of the Delirium Motor Subtyping Scale (DMSS-4). International Psychogeriatrics. 2014;26: 693–702. doi:10.1017/S1041610213002585

15. Kamdar BB, Makhija H, Cotton SA, Fine J, Pollack D, Reyes PA, et al. Development and Evaluation of an Intensive Care Unit Video Series to Educate Staff on Delirium Detection. ATS Sch. 2022;3: 535–547. doi:10.34197/ats-scholar.2022-0011OC

16. Osse RJ, Tulen JHM, Bogers AJJC, Hengeveld MW. Disturbed circadian motor activity patterns in postcardiotomy delirium. Psychiatry Clin Neurosci. 2009;63: 56–64. doi:10.1111/j.1440-1819.2008.01888.x

17. Van Der Kooi AW, Rots ML, Huiskamp G, Klijn FAM, Koek HL, Kluin J, et al. Delirium Detection Based on Monitoring of Blinks and Eye Movements. The American Journal of Geriatric Psychiatry. 2014;22: 1575–1582. doi:10.1016/j.jagp.2014.01.001

18. Gual N, Richardson SJ, Davis DHJ, Bellelli G, Hasemann W, Meagher D, et al. Impairments in balance and mobility identify delirium in patients with comorbid dementia. Int Psychogeriatr. 2019;31: 749–753. doi:10.1017/S1041610218001345

19. Cai L, Gao J, Zhao D. A review of the application of deep learning in medical image classification and segmentation. Ann Transl Med. 2020;8: 713. doi:10.21037/atm.2020.02.44

20. Gabriel P, Rehani P, Troy T, Wyatt T, Choma M, Singh N. Continuous patient monitoring with AI: real-time analysis of video in hospital care settings. Front Imaging. 2025;4. doi:10.3389/fimag.2025.1547166

21. Egnor SER, Branson K. Computational Analysis of Behavior. Annu Rev Neurosci. 2016;39: 217–236. doi:10.1146/annurev-neuro-070815-013845

22. Davoudi A, Malhotra KR, Shickel B, Siegel S, Williams S, Ruppert M, et al. Intelligent ICU for Autonomous Patient Monitoring Using Pervasive Sensing and Deep Learning. Sci Rep. 2019;9: 8020. doi:10.1038/s41598-019-44004-w

23. Raghu R, Nalaie K, Ayala I, Morales Behaine JJ, Garcia-Mendez JP, Friesen H, et al. Harnessing the Power of Technology to Transform Delirium Severity Measurement in the Intensive Care Unit: Protocol for a Prospective Cohort Study. JMIR Res Protoc. 2025;14: e62912. doi:10.2196/62912

24. Gleason A, Richter F, Beller N, Arivazhagan N, Feng R, Holmes E, et al. Detection of neurologic changes in critically ill infants using deep learning on video data: a retrospective single center cohort study. EClinicalMedicine. 2024;78: 102919. doi:10.1016/j.eclinm.2024.102919

25. Collins GS, Moons KGM, Dhiman P, Riley RD, Beam AL, Van Calster B, et al. TRIPOD+AI statement: updated guidance for reporting clinical prediction models that use regression or machine learning methods. BMJ. 2024; e078378. doi:10.1136/bmj-2023-078378

26. Marcantonio ER, Ngo LH, O’Connor M, Jones RN, Crane PK, Metzger ED, et al. 3D-CAM: derivation and validation of a 3-minute diagnostic interview for CAM-defined delirium: a cross-sectional diagnostic test study. Ann Intern Med. 2014;161: 554–561. doi:10.7326/M14-0865

27. Bazarevsky V, Grishchenko I, Raveendran K, Zhu T, Zhang F, Grundmann M. BlazePose: On-device Real-time Body Pose tracking. 2020 [cited 7 Mar 2024]. doi:10.48550/ARXIV.2006.10204

28. Mathis A, Mamidanna P, Cury KM, Abe T, Murthy VN, Mathis MW, et al. DeepLabCut: markerless pose estimation of user-defined body parts with deep learning. Nat Neurosci. 2018;21: 1281–1289. doi:10.1038/s41593-018-0209-y

29. Guetari R, Ayari H, Sakly H. Computer-aided diagnosis systems: a comparative study of classical machine learning versus deep learning-based approaches. Knowl Inf Syst. 2023;65: 3881–3921. doi:10.1007/s10115-023-01894-7

30. Pristyanto Y, Mukarabiman Z, Nugraha AF. Extreme Gradient Boosting Algorithm to Improve Machine Learning Model Performance on Multiclass Imbalanced Dataset. JOIVl1: Int J Inform Visualization. 2023;7: 710–715. doi:10.30630/joiv.7.3.1102

31. Zhao Z, Anand R, Wang M. Maximum Relevance and Minimum Redundancy Feature Selection Methods for a Marketing Machine Learning Platform. 2019 IEEE International Conference on Data Science and Advanced Analytics (DSAA). Washington, DC, USA: IEEE; 2019. pp. 442–452. doi:10.1109/DSAA.2019.00059

32. Ojala M, Garriga GC. Permutation Tests for Studying Classifier Performance. Journal of Machine Learning Research. 2010;11: 1833–1863.

33. Lundberg SM, Lee S-I. A Unified Approach to Interpreting Model Predictions. In: Guyon I, Luxburg UV, Bengio S, Wallach H, Fergus R, Vishwanathan S, et al., editors. Advances in Neural Information Processing Systems. Curran Associates, Inc.; 2017. Available: https://proceedings.neurips.cc/paper_files/paper/2017/file/8a20a8621978632d76c43dfd28b67767-Paper.pdf

34. Bates D, Mächler M, Bolker B, Walker S. Fitting Linear Mixed-Effects Models Using lme4. J Stat Soft. 2015;67. doi:10.18637/jss.v067.i01

35. Genders TSS, Spronk S, Stijnen T, Steyerberg EW, Lesaffre E, Hunink MGM. Methods for calculating sensitivity and specificity of clustered data: a tutorial. Radiology. 2012;265: 910–916. doi:10.1148/radiol.12120509

36. Liu S, Yin Y, Ostadabbas S. In-Bed Pose Estimation: Deep Learning With Shallow Dataset. IEEE Journal of Translational Engineering in Health and Medicine. 2019;7: 1–12. doi:10.1109/JTEHM.2019.2892970

37. Chen K, Gabriel P, Alasfour A, Gong C, Doyle WK, Devinsky O, et al. Patient-Specific Pose Estimation in Clinical Environments. IEEE Journal of Translational Engineering in Health and Medicine. 2018;6: 1–11. doi:10.1109/JTEHM.2018.2875464

38. Avogaro A, Cunico F, Rosenhahn B, Setti F. Markerless human pose estimation for biomedical applications: a survey. Front Comput Sci. 2023;5. doi:10.3389/fcomp.2023.1153160

39. Avogaro A, Cunico F, Rosenhahn B, Setti F. Markerless human pose estimation for biomedical applications: a survey. Front Comput Sci. 2023;5. doi:10.3389/fcomp.2023.1153160

40. Yehoshua R. Chapter 11: Support Vector Machines. 1st ed. Machine Learning Foundations, Volume 1: Supervised Learning. 1st ed. Addison-Wesley; 2025. Available: https://www.oreilly.com/library/view/machine-learning-foundations/9780135337851/

41. Gual N, García-Salmones M, Brítez L, Crespo N, Udina C, Pérez LM, et al. The role of physical exercise and rehabilitation in delirium. Eur Geriatr Med. 2020;11: 83–93. doi:10.1007/s41999-020-00290-6

42. Osse RJ, Tulen JHM, Hengeveld MW, Bogers AJJC. Screening methods for delirium: early diagnosis by means of objective quantification of motor activity patterns using wrist-actigraphy. Interactive CardioVascular and Thoracic Surgery. 2008;8: 344–348. doi:10.1510/icvts.2008.192278

43. Ahmed A, Garcia-Agundez A, Petrovic I, Radaei F, Fife J, Zhou J, et al. Delirium detection using wearable sensors and machine learning in patients with intracerebral hemorrhage. Front Neurol. 2023;14: 1135472. doi:10.3389/fneur.2023.1135472

44. Davoudi A, Malhotra KR, Shickel B, Siegel S, Williams S, Ruppert M, et al. Intelligent ICU for Autonomous Patient Monitoring Using Pervasive Sensing and Deep Learning. Sci Rep. 2019;9: 8020. doi:10.1038/s41598-019-44004-w

45. Richardson S, Murray J, Davis D, Stephan BCM, Robinson L, Brayne C, et al. Delirium and Delirium Severity Predict the Trajectory of the Hierarchical Assessment of Balance and Mobility in Hospitalized Older People: Findings From the DECIDE Study. Newman AB, editor. The Journals of Gerontology: Series A. 2022;77: 531–535. doi:10.1093/gerona/glab081

46. Meagher DJ, O’Hanlon D, O’Mahony E, Casey PR, Trzepacz PT. Relationship Between Symptoms and Motoric Subtype of Delirium. JNP. 2000;12: 51–56. doi:10.1176/jnp.12.1.51

47. Peterson SM, Singh SH, Wang NXR, Rao RPN, Brunton BW. Behavioral and Neural Variability of Naturalistic Arm Movements. eNeuro. 2021;8. doi:10.1523/ENEURO.0007-21.2021

48. Gabriel PG, Chen KJ, Alasfour A, Pailla T, Doyle WK, Devinsky O, et al. Neural correlates of unstructured motor behaviors. J Neural Eng. 2019;16: 066026. doi:10.1088/1741-2552/ab355c

49. Singh SH, Peterson SM, Rao RPN, Brunton BW. Mining naturalistic human behaviors in long-term video and neural recordings. J Neurosci Methods. 2021;358: 109199. doi:10.1016/j.jneumeth.2021.109199

50. Redinbaugh MJ, Saalmann YB. Contributions of Basal Ganglia Circuits to Perception, Attention, and Consciousness. Journal of Cognitive Neuroscience. 2024;36: 1620–1642. doi:10.1162/jocn_a_02177

51. Vardy ERLC, Teodorczuk A, Yarnall AJ. Review of delirium in patients with Parkinson’s disease. J Neurol. 2015;262: 2401–2410. doi:10.1007/s00415-015-7760-1

52. Boord MS, Moezzi B, Davis D, Ross TJ, Coussens S, Psaltis PJ, et al. Investigating how electroencephalogram measures associate with delirium: A systematic review. Clin Neurophysiol. 2021;132: 246–257. doi:10.1016/j.clinph.2020.09.009

53. Bendahan N, Neal O, Ross-White A, Muscedere J, Boyd JG. Relationship Between Near-Infrared Spectroscopy-Derived Cerebral Oxygenation and Delirium in Critically Ill Patients: A Systematic Review. J Intensive Care Med. 2019;34: 514–520. doi:10.1177/0885066618807399

54. Yokota H, Ogawa S, Kurokawa A, Yamamoto Y. Regional cerebral blood flow in delirium patients. Psychiatry and Clinical Neurosciences. 2003;57: 337–339. doi:10.1046/j.1440-1819.2003.01126.x

55. Fleischmann R, Andrasch T, Warwas S, Kunz R, Gross S, Witt C, et al. Predictors of post-stroke delirium incidence and duration: Results of a prospective observational study using high-frequency delirium screening. Int J Stroke. 2023;18: 278–284. doi:10.1177/17474930221109353

56. Miyamoto E, Tomimoto H, Nakao Si S, Wakita H, Akiguchi I, Miyamoto K, et al. Caudoputamen is damaged by hypocapnia during mechanical ventilation in a rat model of chronic cerebral hypoperfusion. Stroke. 2001;32: 2920–2925. doi:10.1161/hs1201.100216

57. Lindroth H, Nalaie K, Raghu R, Ayala IN, Busch C, Bhattacharyya A, et al. Applied Artificial Intelligence in Healthcare: A Review of Computer Vision Technology Application in Hospital Settings. Journal of Imaging. 2024;10. doi:10.3390/jimaging10040081

58. Parker S, Gilstrap D, Bedoya A, Lee P, Deshpande K, Gabriel PG, et al. Continuous Artificial Intelligence Video Monitoring of ICU Patient Activity for Detecting Sedation, Delirium and Agitation. C35 TOPICS IN CRITICAL CARE AND RESPIRATORY FAILURE. American Thoracic Society; 2022. pp. A5719–A5719. doi:10.1164/ajrccm-conference.2022.205.1_MeetingAbstracts.A5719

59. Tatum WO, Mani J, Jin K, Halford JJ, Gloss D, Fahoum F, et al. Minimum standards for inpatient long-term video-EEG monitoring: A clinical practice guideline of the international league against epilepsy and international federation of clinical neurophysiology. Clinical Neurophysiology. 2022;134: 111–128. doi:10.1016/j.clinph.2021.07.016

60. Kane GA, Lopes G, Saunders JL, Mathis A, Mathis MW. Real-time, low-latency closed-loop feedback using markerless posture tracking. Berman GJ, Behrens TE, Berman GJ, Branco T, editors. eLife. 2020;9: e61909. doi:10.7554/eLife.61909

61. Kimchi EY, Neelagiri A, Whitt W, Sagi AR, Ryan SL, Gadbois G, et al. Clinical EEG slowing correlates with delirium severity and predicts poor clinical outcomes. Neurology. 2019;93. doi:10.1212/WNL.0000000000008164

62. Contreras M, Kapoor S, Zhang J, Davidson A, Ren Y, Guan Z, et al. A large language model for delirium prediction in the intensive care unit using structured electronic health records. Scientific Reports. 2025;15: 38890. doi:10.1038/s41598-025-22634-7

63. O’Keeffe ST, Lavan JN. Clinical significance of delirium subtypes in older people. Age Ageing. 1999;28: 115–119. doi:10.1093/ageing/28.2.115

64. Emekli E, Hasanli J, Alici Y. Missed Delirium Diagnoses in Elderly Inpatients: Factors Associated with Diagnostic Inaccuracy by Non-Psychiatrist Physicians. PBS. 2025;15: 172. doi:10.5455/PBS.20240927090147

65. Hshieh TT, Yang T, Gartaganis SL, Yue J, Inouye SK. Hospital Elder Life Program: Systematic Review and Meta-analysis of Effectiveness. Am J Geriatr Psychiatry. 2018;26: 1015–1033.

66. Kim MS, Rhim HC, Park A, Kim H, Han K-M, Patkar AA, et al. Comparative efficacy and acceptability of pharmacological interventions for the treatment and prevention of delirium: A systematic review and network meta-analysis. J Psychiatr Res. 2020;125: 164–176. doi:10.1016/j.jpsychires.2020.03.012

