## Supplement for "Video-based Detection of Delirium in Hospitalized Adults"

### Supplementary Methods

**Deep Learning Model Training and Testing Specifications**

DeepLabCut was utilized to fine-tune the ResNet101 model for human pose estimation on the patient video dataset. The input image size was 1920x1440 pixels, and the output was 35 pairs of coordinates corresponding to each keypoint. The algorithm was fine-tuned for 700,000 iterations with default parameters.

Training Parameters

- Training Fraction: 0.8 (Proportion of dataset split into training)
- dot_size: 3 (point size)
- batch_size: 8
- optimizer: sgd (stochastic gradient descent)
- lr_init: 0.0005 (initial learning rate)
- max_iters: 789,000 (maximum iterations)
- multi_step: (learning rate schedule)
  - [0.005, 10000]
  - [0.02, 430000]
  - [0.002, 730000]
  - [0.001, 1030000]
- init_weights: resnet101 (initial weights)
- scale_jitter_lo: 0.5
- scale_jitter_up: 1.25 (each image was rescaled within range of jitter values to augment training)
- p-cutoff: 0.812 (likelihood threshold for visibility)

The algorithm was evaluated on a held-out testing dataset, comprising of 20% of the total dataset. Model performance was assessed using Euclidean distance for human labeled points that were identified by the model with a sufficient confidence threshold. Euclidean distance was defined as $\sqrt{{(x_{m}-x_{g})}^{2}+{(y_{m}-y_{g})}^{2}}$ where $(x_{g},y_{g})$ are the ground truth labels and $(x_{m},y_{m})$ are the machine-labeled points. The confidence threshold for the model (p-cutoff) was selected to optimize the F1 score, which is the harmonic mean of precision and recall of point detection on the training dataset.

**Deep Learning Model Benchmarking**

The fine-tuned model termed “DLC” was evaluated against FaceMesh and BlazePose from the MediaPipe suite. The FaceMesh and BlazePose models were trained on datasets of 30k mobile camera photos of people-in-the-wild and 85k photos of people performing common poses or fitness exercises, respectively. These models were used as is and not fine-tuned on our video cohort. Model performance was compared by using 33 shared points that were identified by FaceMesh, BlazePose, and DLC. The conversion between DLC, BlazePose, and FaceMesh points is detailed in the data_key.csv file.

**Feature Pre-Processing and Selection**

Given the large number of behavioral features calculated based on the video data, feature selection was performed to improve model fit. In addition to using all features, different feature selection methods were considered: chi-square based univariate selection, lasso regression, mutual information (MI), and minimum redundancy maximum relevancy (mRMR) [27]. Additionally, the number of features selected (k) was varied between 10, 20, 40, and 80 for the chi-square, MI, and mRMR-based methods.

For chi-square based feature selection, the feature data was min-max scaled to ensure it was non-negative before performing feature selection using the SelectKBest function in scikit-learn. Lasso regression was implemented on the feature data using the LassoCV linear model where features with non-zero coefficients were selected. mRMR was implemented using the mrmr-selection package. Mutual Information was implemented using the *mutual_info* function from scikit-learn.

**Delirium Classification Models**

Four different machine learning models were fit: support vector machine, logistic regression, extreme gradient boosting [26], and random forest-based models. Support vector machine, Logistic regression, and the Random Forest model were implemented using the *LinearSVC*, *LogisticRegression,* and *RandomForestClassifier* functions from the sci-kit learn library, respectively. The Gradient Boosting model was implemented using the *XGBClassifier* function from the xgboost library.

**Missing Values**

Behavioral features with missing values were imputed using zero substitution. Behavioral features with infinite values, which can arise from ratios when the denominator approaches zero, were replaced with the maximum or minimum finite value observed in that feature across the training dataset. This approach was applied uniformly prior to any feature selection or model fitting.

**Considerations for Future Implementation**

The quality of video data (lighting, occlusion, etc.) and its effect on model predictions was not formally evaluated. Future work should define protocols for handling poor quality input video data. User interaction is required in three phases: video capture, video screening, and interpretation of model results. The camera should be mounted at the foot of the hospital bed as described in Methods: Video Data Collection. Prior to being used in the deep learning model, the videos were manually screened to ensure they did not contain fully masked patients, head wraps from epilepsy monitoring, or other video file errors. Future research could automate this process of screening videos. The output of the model is intended to serve as a screening tool rather than a standalone clinical diagnosis and should be interpreted in the context of clinical findings by a trained healthcare professional.

### Supplementary Figures

Supplementary Figure 1**.** Analysis Workflow

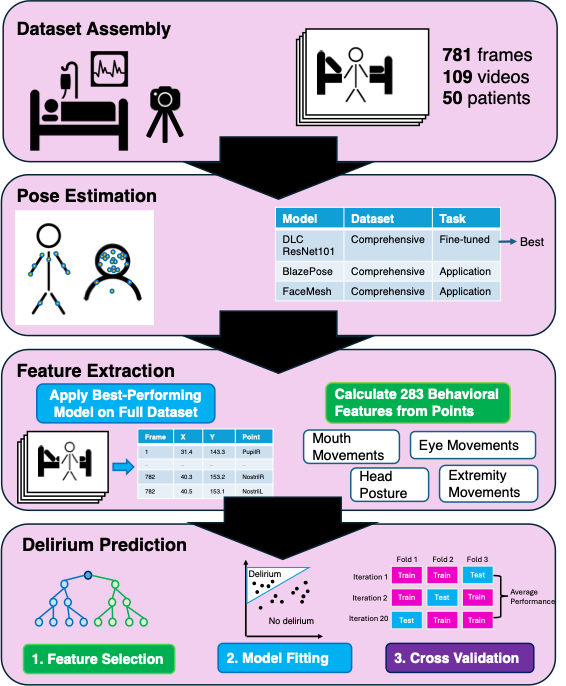

Supplementary Figure 2**.** Assessment of repeated measures, showing how many participants contributed each number of videos per participant in the comprehensive dataset.

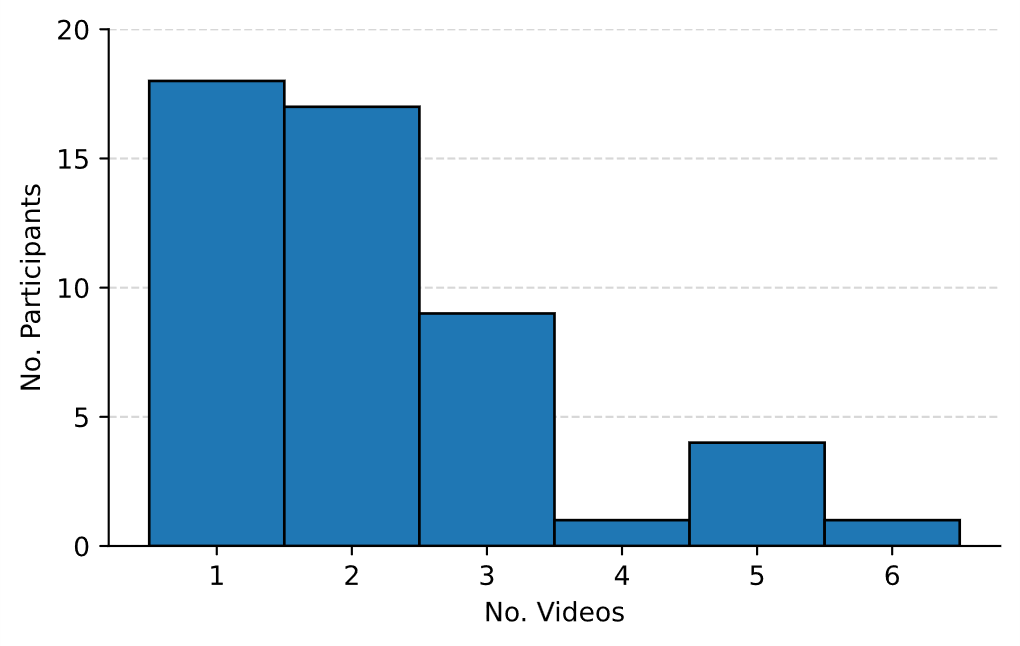

Supplementary Figure 3**.** Permutation Analyses for Different Machine Learning Models

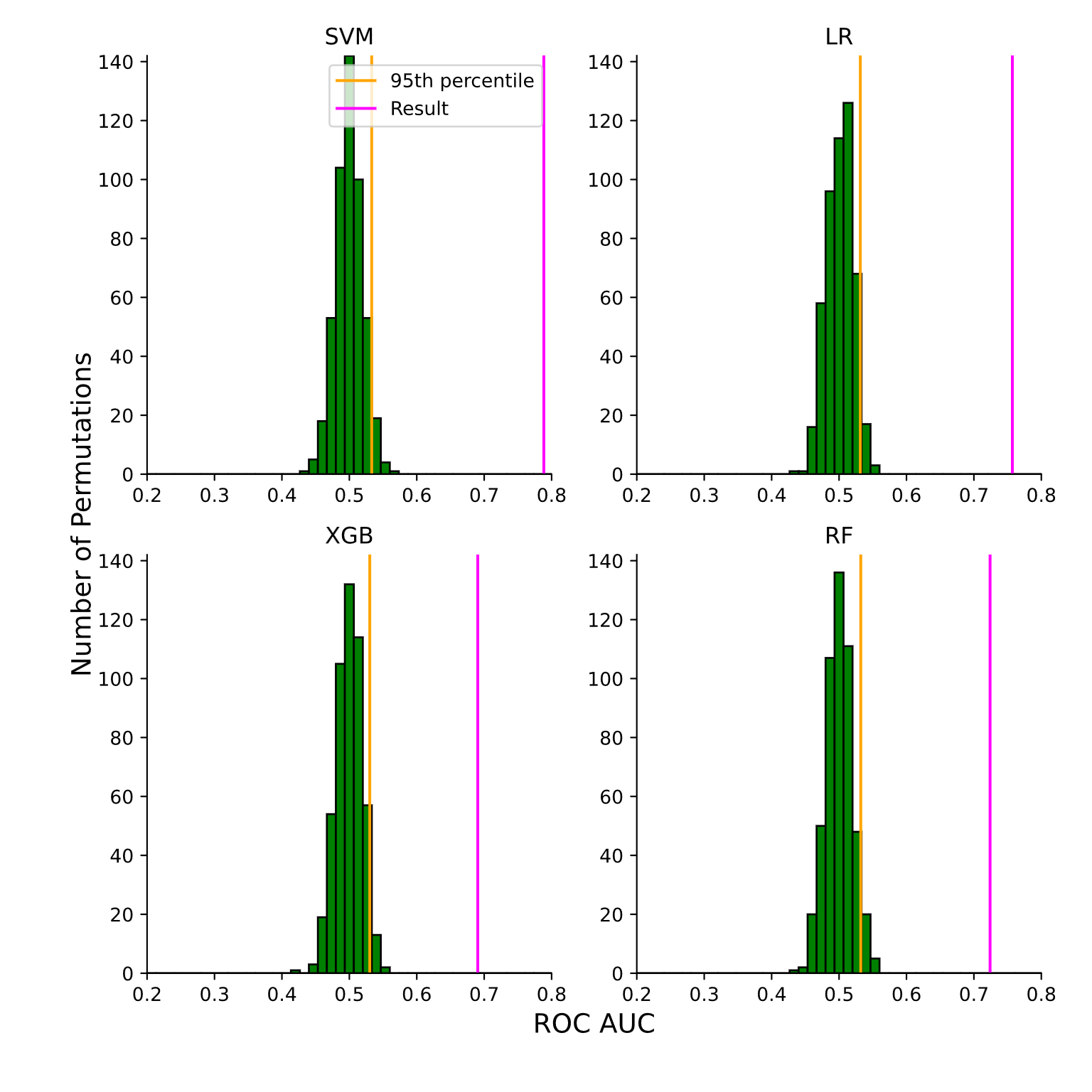

Models were fitted on a shuffled version of the video dataset for 500 trials using the same feature selection and model fitting procedure. The 95^th^ percentile of the null distribution is denoted by the orange line. Estimated model performance is plotted using the pink line for support vector machine (SVM), logistic regression (LR), extreme gradient boosting (XGB), and random forest (RF) algorithms

Supplementary Figure 4**.** Feature Importance for Different Machine Learning Models

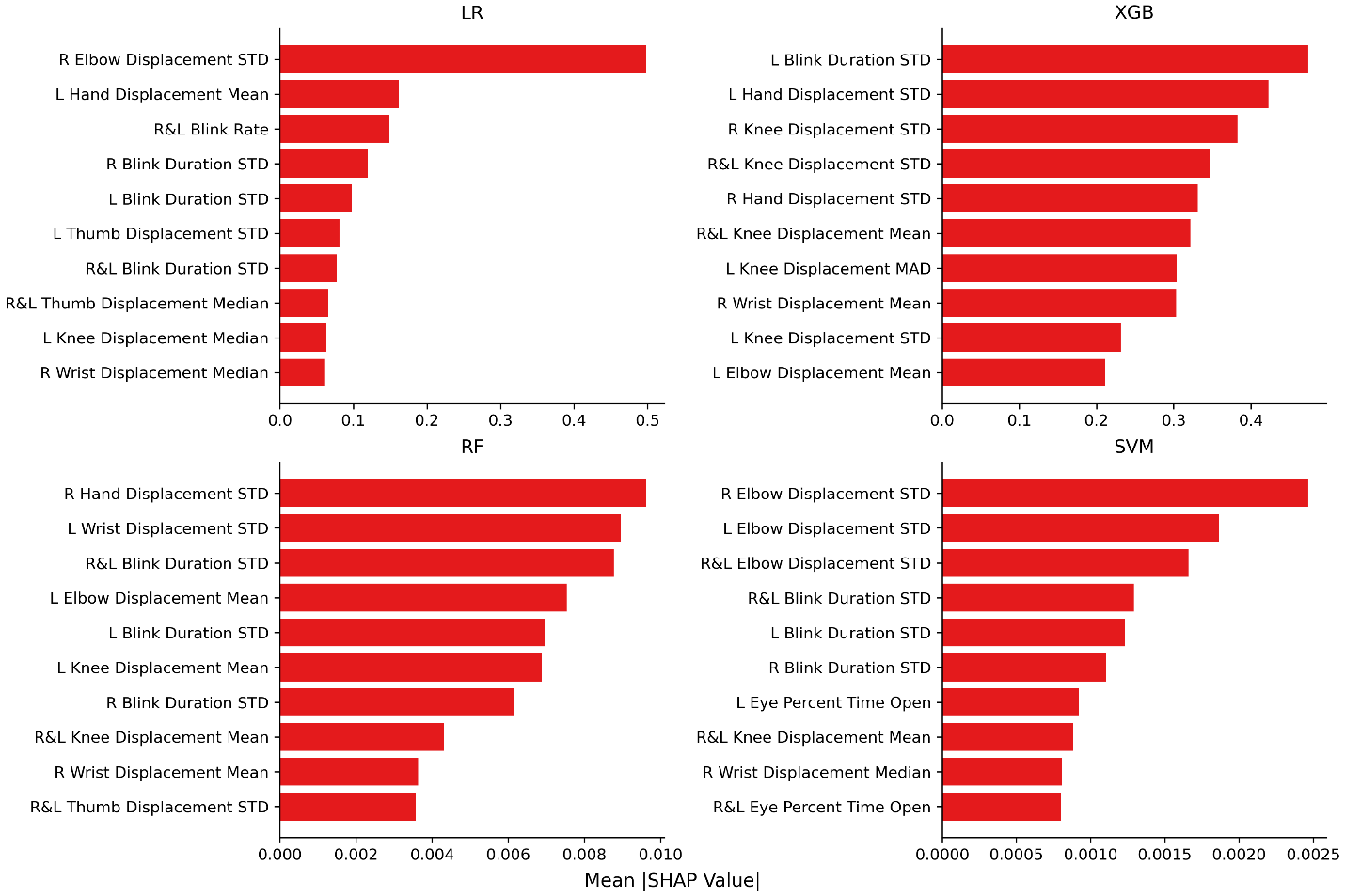
The top 10 features used in each algorithm are ranked in descending order of importance by their mean absolute Shapley analysis (SHAP) values across folds and repetitions for the best combination of algorithm and feature selection method. Features of the highest importance clustered around the upper extremities and eyes. Displacement was measured as the frame-by-frame change in position for the keypoints, with summary measures being taken across all changes between frames. “R” denotes right, “L” denotes left, and “R&L” denotes that the metric was averaged across both sides to calculate this feature. “STD” denotes standard deviation, while “MAD” denotes mean absolute deviation. See Supplementary Table 4 for further descriptions of how each feature was calculated.

### Supplementary Tables

Supplementary Table 1 Keypoints labeled for video frames

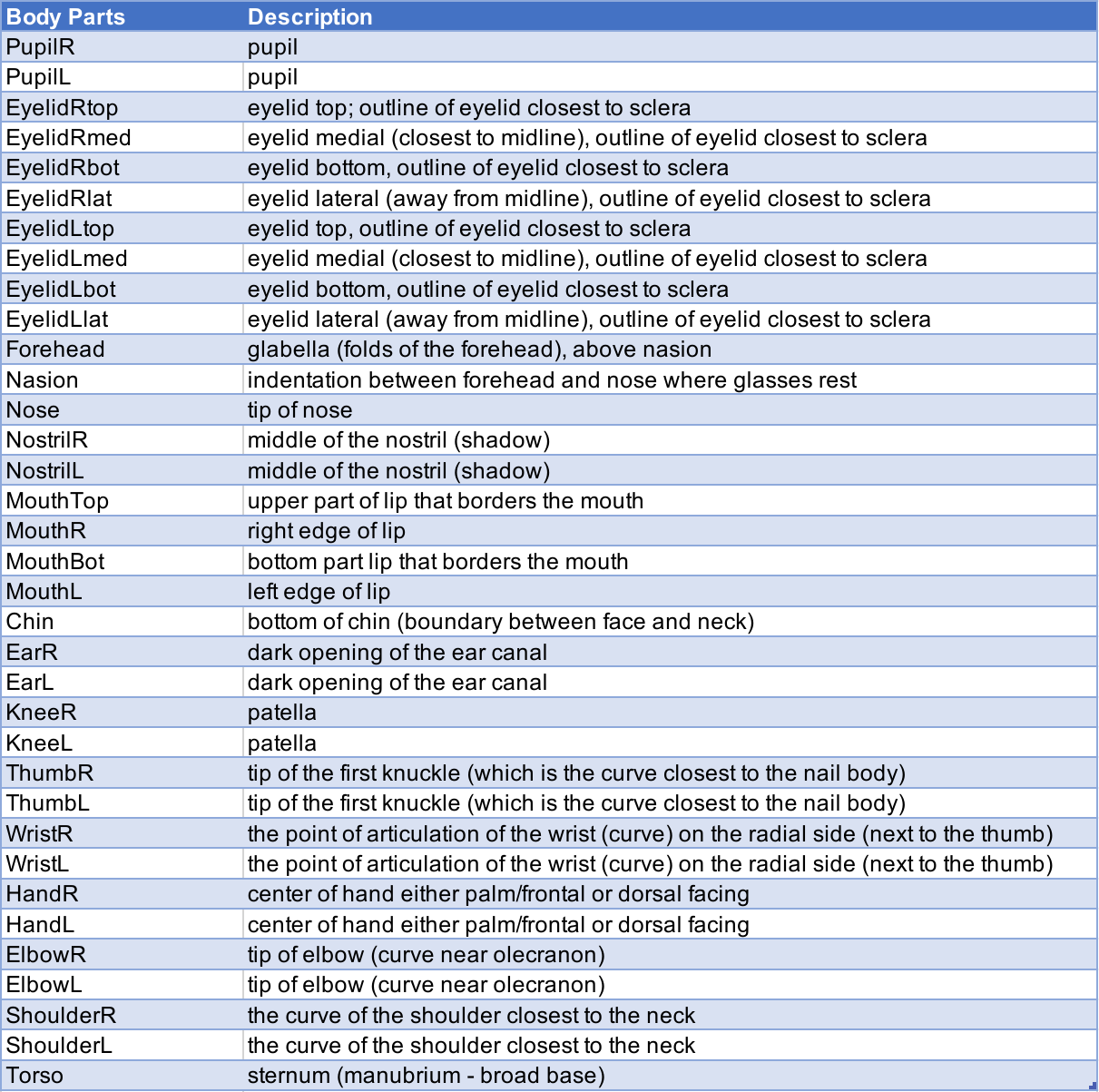

Supplementary Table 2 Composition of Restricted and Comprehensive Model Datasets

**Initial/Restricted Model Dataset:**

|  | **Total** | **Training** | **Testing** | **Validation** |
| --- | --- | --- | --- | --- |
| # frames | 400 | 251 | 63 | 86 |
| # videos | 45 | 35 | 25 | 10 |
| # patients | 40 | 35 | 25 | 10 |

**Comprehensive Model Dataset:**

|  | **Total** | **Training** | **Testing** |
| --- | --- | --- | --- |
| # frames | 782 | 625 | 157 |
| # videos | 109 | 109 | 81 |
| # patients | 50 | 50 | 44 |

The Total column represents the total numbers of labeled frames in the Initial Restricted or Subsequent Comprehensive datasets. Frames were split into Training and Testing sets for the Comprehensive dataset. The Restricted dataset also included a Validation set with videos not included in Training or Testing sets.

Supplementary Table 3 Video Feature Domains

| **Domain** | **Features** | **Number** |
| --- | --- | --- |
| Eye movement | Pupil movements, horizontal pupil movements, vertical pupil movements, horizontal eyelid axis, vertical eyelid axis | 15 |
| Blinks | Blink rate, blink duration, eye aspect ratio, % of frames with eyes open | 7 |
| Head movements | Roll, pitch, yaw, and nose aspect ratio | 4 |
| Mouth movements | % of frames with mouth open, mouth vertical distance, mouth horizontal distance, upper lip movement, and lower lip movement | 6 |
| Upper extremity | % of frames with hands higher than elbow, % of frames with hands higher than chin, wrist displacement, hand displacement, elbow movement, shoulder movement, arm movements (wrist-shoulder-elbow angle) | 23 |
| Lower extremity | Knee movement | 3 |
| Summary Statistics | mean, median, standard deviation, and median absolute deviation | X 4 |
| Total |  | 232 |

Supplementary Table 4. Behavioral Feature Descriptions

| **Feature Names** | **Variable Names** | **Calculation** | **Notes** |
| --- | --- | --- | --- |
| Percent eyes open, Percent right eye open, Percent left eye open | - perc_eye_open - R_eyes_open - L_eyes_open | % of frames where at least one pupil is detected, % of frames where right pupil is detected, % of frames where left pupil is detected |  |
| Percent mouth open | - per_mouth_open | % of frames where the mouth vertical distance was greater than the mouth horizontal distance times .3 |  |
| Mean upper lip movement, mean lower lip movement, average mean lip movement, median upper lip movement, median lower lip movement, average median lip movement, upper lip movement variability, lower lip movement variability, average lip movement variability, upper lip movement median absolute deviation, lower lip movement median absolute deviation, average lip movement median absolute deviation | - mean_Tmouthdiff - mean_Bmouthdiff - Avg_mean_mouthdiff - median_Tmouthdiff - median_Bmouthdiff - Avg_median_mouthdiff - std_Tmouthdiff - std_Bmouthdiff - Avg_std_mouthdiff - MAD_Tmouthdiff - MAD_Bmouthdiff - avg_MAD_mouthdiff | Upper lip distance was calculated as the difference in position between consecutive frames, lower lip distance was calculated as the difference in position between consecutive frames |  |
| Mean right pupil movement, mean left pupil movement, etc. | - mean_Rpupildist, - mean_Lpupildist, - Avg_mean_pupildist - median_Rpupildist - median_Lpupildist - Avg_median_pupildist - std_Rpupildist - std_Lpupildist - Avg_std_pupildist - MAD_Rpupildist | Right pupil movement was calculated as the distance between consecutive frames, left pupil movement was calculated as the distance between consecutive frames |  |
| Mean right vertical pupil movements, mean left vertical pupil movements, etc. | - mean_RVeyemvmt, - mean_LVeyemvmt, - avg_mean_Veyemvmt, - median_RVeyemvmt, - median_LVeyemvmt, - avg_median_Veyemvmt, - std_RVeyemvmt, - std_LVeyemvmt, - avg_std_Veyemvmt, - MAD_RVeyemvmt, - MAD_LVeyemvmt, - avg_MAD_Veyemvmt | 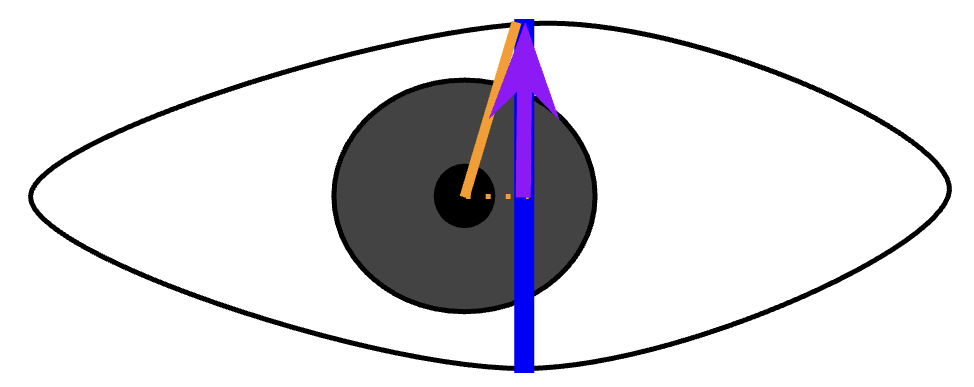  $\frac{pupilR-eyelidTop \cdot vertical\_eyelid\_axis}{\parallel vertical\_eyelid\_axis\parallel}$  Right vertical pupil movement is represented by the purple vector |  |
| Mean right horizontal pupil movement, mean left horizontal pupil movements, etc. | - mean_RHeyemvmt, - mean_LHeyemvmt, - avg_mean_Heyemvmt, - median_RHeyemvmt - median_LHeyemvmt, - avg_median_Heyemvmt, - std_RHeyemvmt, - std_LHeyemvmt, - avg_std_Heyemvmt, - MAD_RHeyemvmt, - MAD_LHeyemvmt, - avg_MAD_Heyemvmt | 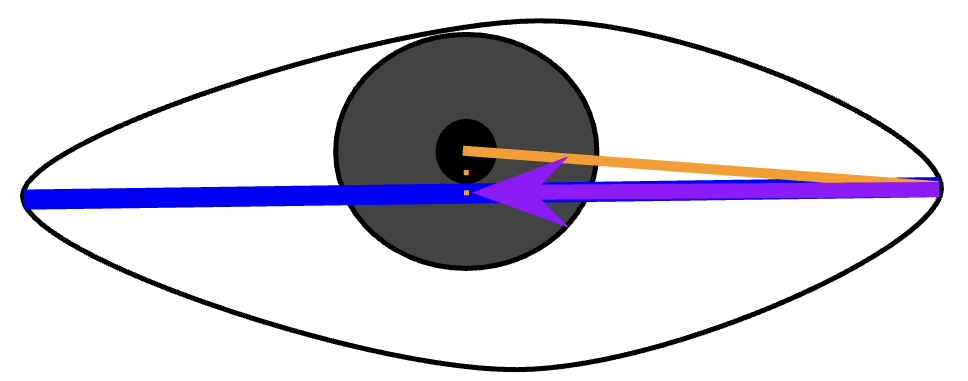  $\frac{pupilL-eyelidmed \cdot horizontal\_eyelid\_axis}{\parallel horizontal\_eyelid\_axis\parallel}$  Left horizontal pupil movement is represented by the purple vector |  |
| Mean right horizontal eyelid axis length, mean left horizontal eyelid axis length, etc. | - mean_RHeyelidaxis, - mean_LHeyelidaxis, - avg_mean_Heyelidaxis, - median_RHeyelidaxis, - median_LHeyelidaxis, - avg_median_Heyelidaxis, - std_RHeyelidaxis, - std_LHeyelidaxis, - avg_std_Heyelidaxis, - MAD_RHeyelidaxis, - MAD_LHeyelidaxis, - avg_MAD_Heyelidaxis | 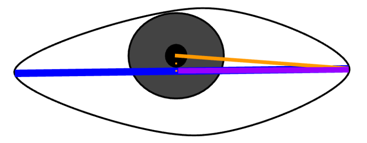  The horizontal right eyelid axis is represented by the blue vector |  |
| Mean right vertical eyelid axis length, mean left vertical eyelid axis length, etc. | - mean_RVeyelidaxis - mean_LVeyelidaxis - avg_mean_Veyelidaxis - median_RVeyelidaxis - median_LVeyelidaxis - avg_median_Veyelidaxis - std_RVeyelidaxis - std_LVeyelidaxis - avg_std_Veyelidaxis - MAD_RVeyelidaxis - MAD_LVeyelidaxis - avg_MAD_Veyelidaxis | 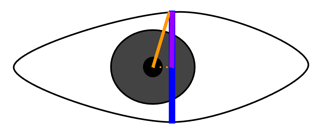  The vertical right eyelid axis is represented by the blue vector |  |
| Mean right eye aspect ratio, mean left eye aspect ratio, etc. | - mean_R_EAR - mean_L_EAR - avg_mean_EAR - median_R_EAR - median_L_EAR - avg_median_EAR - std_R_EAR - std_L_EAR - avg_std_EAR - MAD_R_EAR - MAD_L_EAR - avg_MAD_EAR | 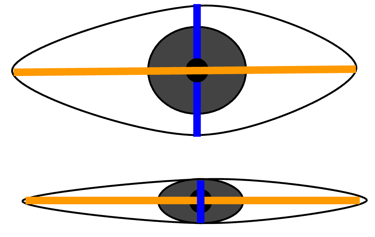    Eye aspect ratio helps gauge eye openness and is calculated as the ratio of vertical eyelid distance and horizontal eyelid distance.  $\frac{\parallel vertical\_eyelid\_axis\parallel}{\parallel horizontal\_eyelid\_axis\parallel}$ | Rosebrock, A. (2017, April 24). Eye blink detection with OpenCV, Python, and dlib. *Pyimagesearch*. <https://pyimagesearch.com/2017/04/24/eye-blink-detection-opencv-python-dlib/> |
| Mean vertical mouth distance, mean horizontal mouth distance, etc. | - mean_Vmouthdist - mean_Hmouthdist - avg_mean_mouthdist - median_Vmouthdist - median_Hmouthdist - avg_median_mouthdist - std_Vmouthdist - std_Hmouthdist - avg_std_mouthdist - MAD_Vmouthdist - MAD_Hmouthdist - avg_MAD_mouthdist | Vertical mouth distance is calculated as the distance between the upper and lower lip points. Horizontal mouth distance is calculated as the distance between the left and right mouth points. |  |
| Mean right blink duration, mean left blink duration, average blink duration, etc. | - mean_Rblinkt - mean_Lblinkt - avg_mean_blinkt - median_Rblinkt - median_Lblinkt - avg_median_blinkt - std_Rblinkt - std_Lblinkt - avg_std_blinkt - MAD_Rblinkt - MAD_Lblinkt - avg_MAD_blinkt | Blink duration is calculated by examining the number of frames between eyes open-to-close and close-to-open events. Eyes were defined as close when the pupil was not detected. |  |
| Right eye blink rate, left eye blink rate, average blink rate | - Rblink_rate - Lblink_rate - avg_blink_rate | Blink rate is calculated as the number of blinks per minute. Blinks are defined based on pupil detection. Blink rate was specifically equal to the number of eyes open-to-close events divided by the frame rate and multiple by 60. |  |
| Percent of frames where the right hand is higher than the right elbow, percent of frames where the left hand is higher than the left elbow, percent of frames where the right hand is higher than the chin, percent of frames where the left hand is higher than the chin | - Rhandhigher_elb - Lhandhigher_elb, - Rhandhigher_chin - Lhandhigher_chin | The % of frames where the hand is higher than the elbow is calculated by comparing the y coordinate of the hand and elbow points in frames where both were detected. This is a rough proxy for carphology.  The % of frames where the hand is higher than the chin is calculated by comparing the y coordinate of the hand and chin points in frames where both were detected. This is a rough proxy for carphology. |  |
| Roll | - mean_roll - median_roll - std_roll - MAD_roll | Roll head pose captures rotation in-plane movements. Roll was calculated as $\arctan\left( \frac{y_{1}}{x_{1}} \right)$  where y_1_ is the difference in the y coordinates between the left and right eye midpoints and x_1_ is the difference in the x coordinates between the left and right eye midpoints. Roll angle is an “estimate of the inclination angle on the roll axis” and is defined with respect to current eye position here (Arcoverde et al., 2014)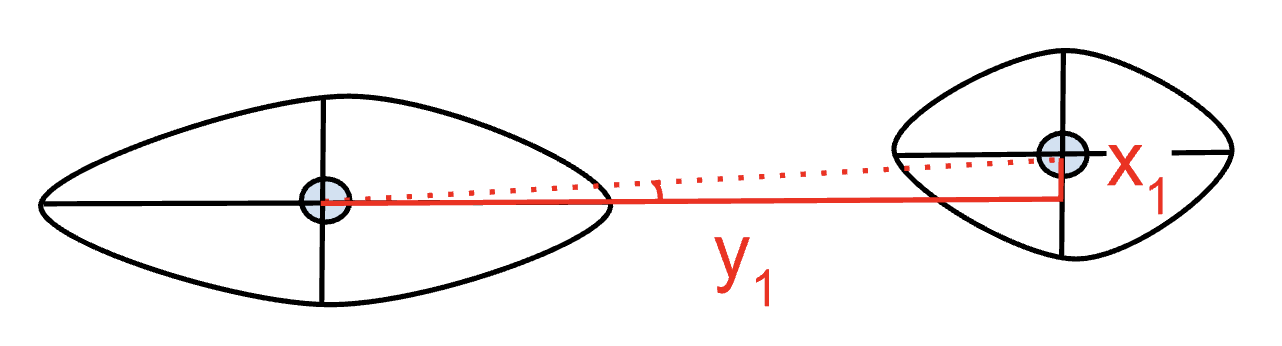 | Arcoverde Neto, E. N., Duarte, R. M., Barreto, R. M., Magalhães, J. P., Bastos, C. C. M., Ren, T. I., & Cavalcanti, G. D. C. (2014). Enhanced real-time head pose estimation system for mobile device. *Integrated Computer-Aided Engineering*, *21*(3), 281–293. <https://doi.org/10.3233/ICA-140462> |
| Pitch | - mean_pitch - median_pitch - std_pitch - MAD_pitch | Pitch head pose captures head up and down movements. Pitch was calculated as the differences in the nose y coordinates between consecutive frames. | Arcoverde Neto, E. N., Duarte, R. M., Barreto, R. M., Magalhães, J. P., Bastos, C. C. M., Ren, T. I., & Cavalcanti, G. D. C. (2014). Enhanced real-time head pose estimation system for mobile device. *Integrated Computer-Aided Engineering*, *21*(3), 281–293. <https://doi.org/10.3233/ICA-140462> |
| Yaw | - mean_yaw - median_yaw - std_yaw - MAD_yaw | Yaw head pose captures head side to side movements. Yaw was calculated as the differences in nose x coordinate between consecutive frames. | Arcoverde Neto, E. N., Duarte, R. M., Barreto, R. M., Magalhães, J. P., Bastos, C. C. M., Ren, T. I., & Cavalcanti, G. D. C. (2014). Enhanced real-time head pose estimation system for mobile device. *Integrated Computer-Aided Engineering*, *21*(3), 281–293. <https://doi.org/10.3233/ICA-140462> |
| Nose Aspect Ratio | - mean_NAR - median_NAR - std_NAR - MAD_NAR | 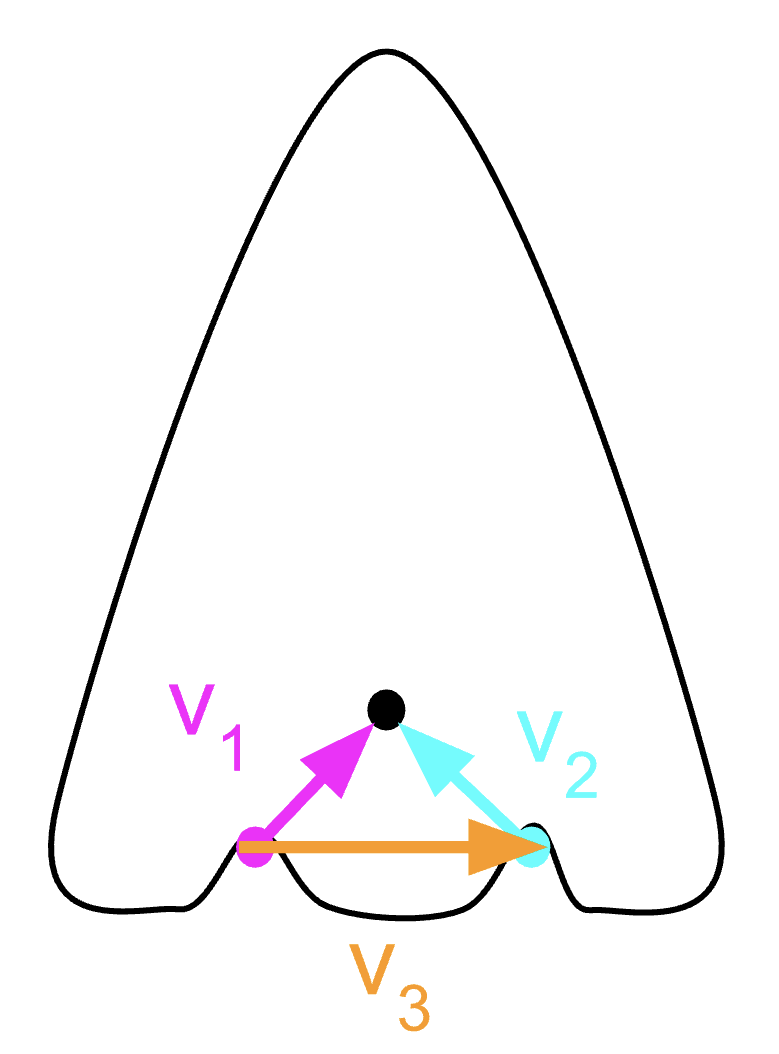  Nose aspect ratio was another parameters used to represent in-plane head rotation. It was defined as  $\frac{v_{1}+v_{2}}{2*{\parallel v}_{3}\parallel}$ |  |
| Mean right thumb displacement, mean left thumb displacement, average mean thumb displacement, median right thumb displacement, median left thumb displacement, average median thumb displacement, right thumb displacement standard deviation, left thumb displacement standard deviation, etc. | - mean_thumbR - mean_thumbL - avg_mean_thumb - median_thumbR - median_thumbL - avg_median_thumb - std_thumbR - std_thumbL - avg_std_thumb - MAD_thumbR - MAD_thumbL - avg_MAD_thumb | Thumb displacement was calculated frame-to-frame by computing the Euclidean distance between consecutive (x,y) coordinates of the tip of the thumb |  |
| Mean right hand displacement, mean left hand displacement, average hand displacement, median right hand displacement, median left hand displacement, average median hand displacement, right hand displacement standard deviation, left hand displacement standard deviation, etc. | - mean_handR - mean_handL - avg_mean_hand - median_handR - median_handL - avg_median_hand - std_handR - std_handL - avg_std_hand - MAD_handR - MAD_handL - avg_MAD_hand | Hand displacement was calculated frame-to-frame by computing the Euclidean distance between consecutive (x,y) coordinates of the center of each hand |  |
| Mean right wrist displacement, mean left wrist displacement, average wrist displacement, median right wrist displacement, median left wrist displacement, average median wrist displacement, right wrist displacement standard deviation, left wrist displacement standard deviation, etc. | - mean_wristR - mean_wristL - avg_mean_wrist - median_wristR - median_wristL - avg_median_wrist - std_wristR - std_wristL - avg_std_wrist - MAD_wristR - MAD_wristL - avg_MAD_wrist | Wrist displacement was calculated frame-to-frame by computing the Euclidean distance between consecutive (x,y) coordinates of the point of the wrist that articulates with the radius |  |
| Mean right elbow displacement, mean left elbow displacement, average elbow displacement, median right elbow displacement, median left elbow displacement, average median elbow displacement, right elbow displacement standard deviation, left elbow displacement standard deviation, etc. | - mean_elbowR - mean_elbowL - avg_mean_elbow - median_elbowR - median_elbowL - avg_median_elbow - std_elbowR - std_elbowL - avg_std_elbow - MAD_elbowR - MAD_elbowL - avg_MAD_elbow | Elbow displacement was calculated frame-to-frame by computing the Euclidean distance between consecutive (x,y) coordinates of the tip of the elbow |  |
| Shoulder R-Shoulder L-Torso Triangle (STT or chest openness) | - mean_STT - median_STT - std_STT - MAD_STT | $\frac{\left\vert\left\vert Torso-Shoulder_{R} \right\vert\right\vert+\left\vert\left\vert Torso-Shoulder_{L} \right\vert\right\vert}{2\cdot\left\vert\left\vert Shoulder_{R} - Shoulder_{L} \right\vert\right\vert}$  The euclidean distance between each shoulder and torso was calculated for each frame and normalized by dividing two times the shoulder width. This metric represents the relative expansion of the upper body. Higher values indicate greater chest openess. | 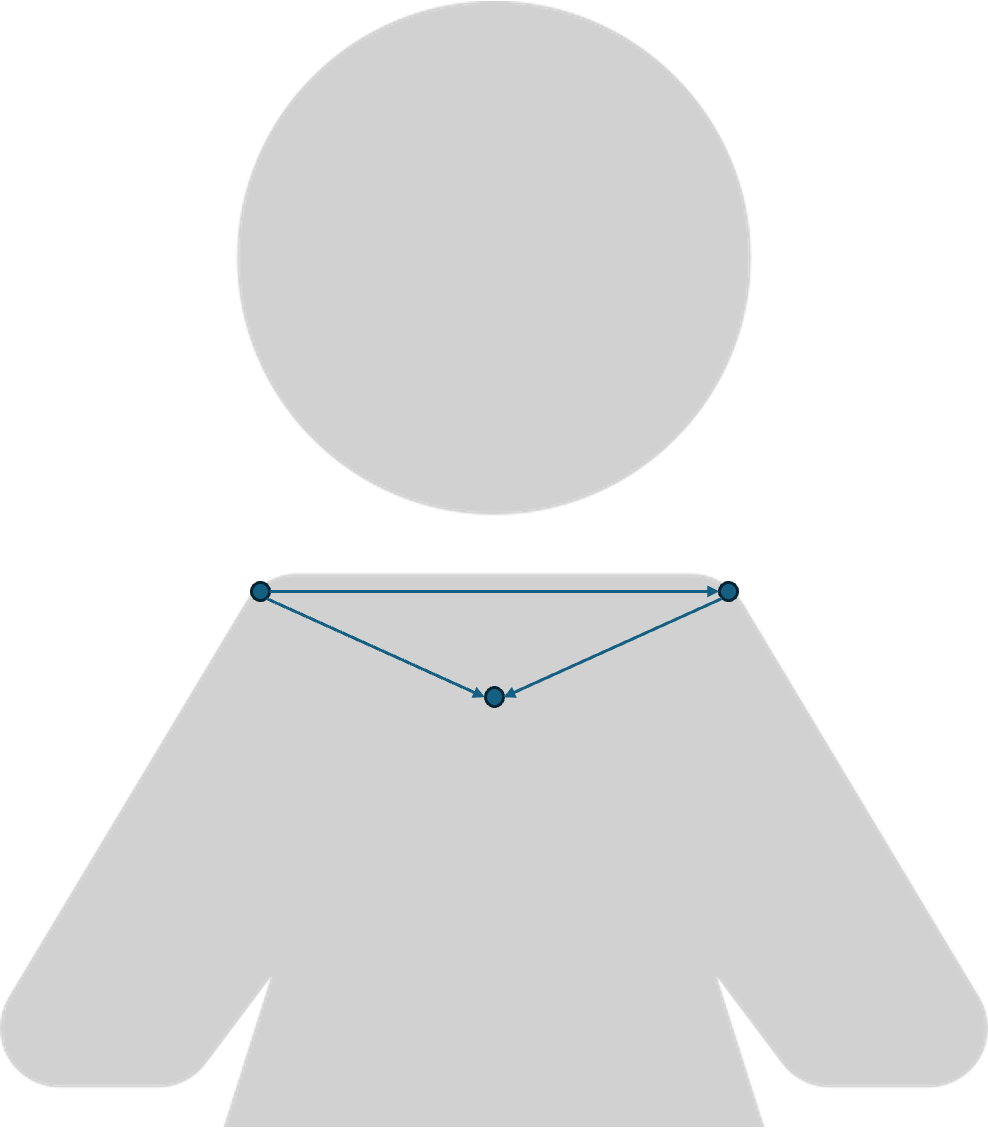 |
| Shoulder-Wrist-Elbow Angle (SWE) | - mean_sweR - mean_swe - avg_mean_swe - median_sweR - median_sweL - avg_median_swe - std_sweR - std_sweL - avg_std_swe - MAD_sweR - MAD_sweL - avg_MAD_swe | The shoulder-wrist-elbow angle is calculated for each frame by computing the arctangent of the ratio between shoulder-elbow length (upper arm) and elbow-wrist (forearm) length. It approximates changes in arm configuration. Note that this does not represent a true anatomical joint angle. | 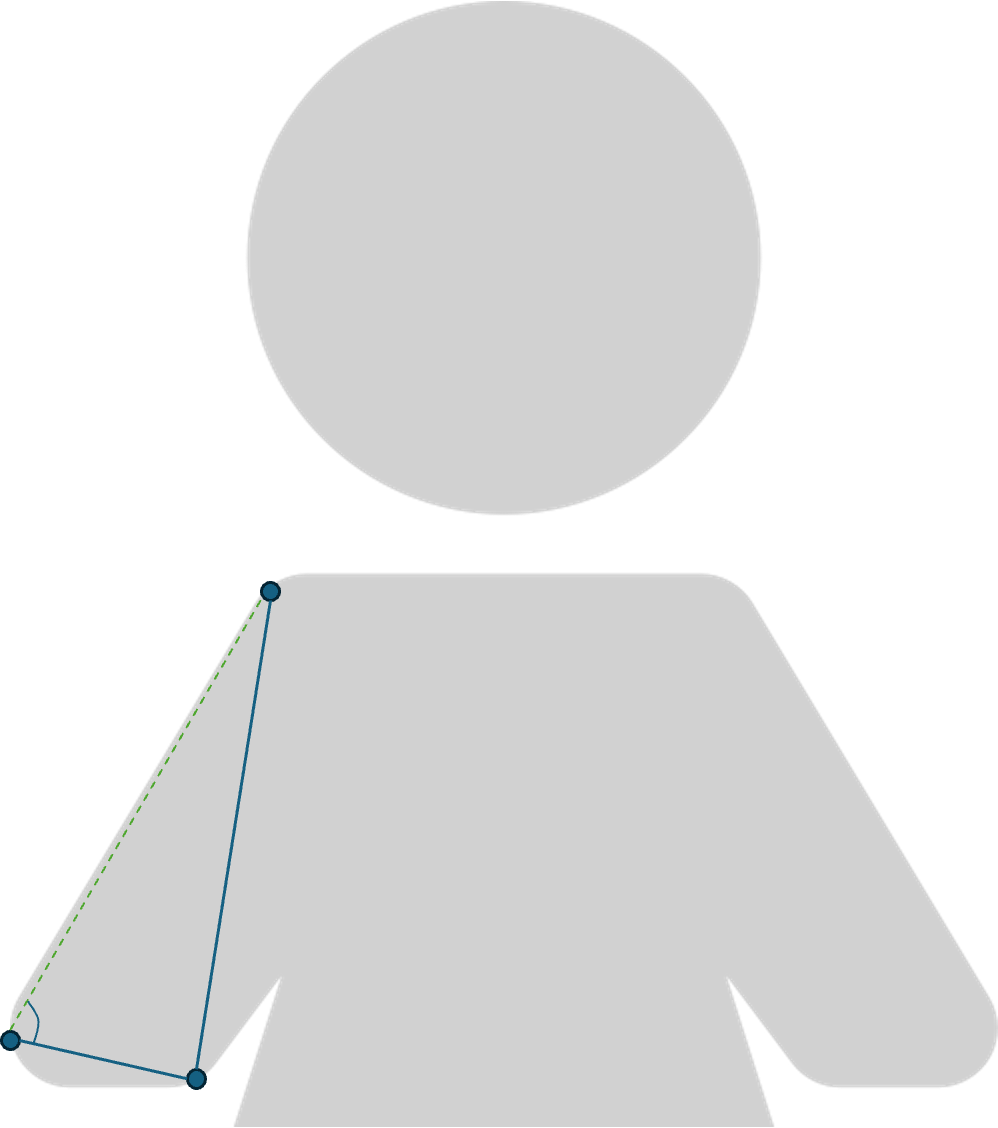 |
| Wrist-Shoulder-Elbow Angle (WSE) | mean_wseR, mean_wseL, avg_mean_wse, median_wseR, median_wseL, avg_median_wse, std_wseR, std_wseL, avg_std_wse, MAD_wseR, MAD_wseL, avg_MAD_wse | The wrist-shoulder-elbow angle is calculated for each frame by computing the arctangent of the ratio between the elbow-wrist length (forearm) and shoulder-elbow length (upper arm). Note that this does not represent a true anatomical joint angle. | 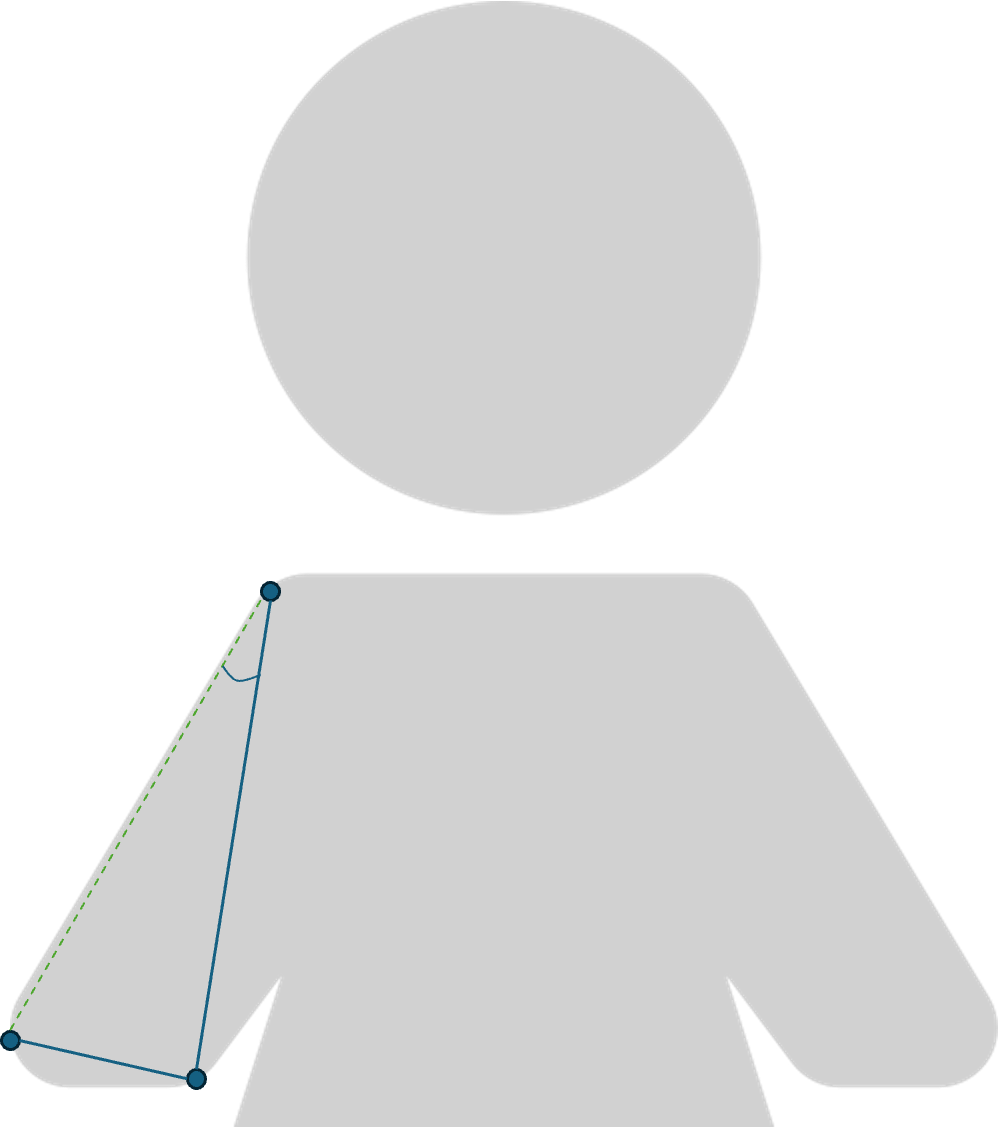 |
| Thumb-Wrist-Hand Angle (TWH) | mean_twhR, mean_twhL, avg_mean_twh, median_twhR, median_twhL, avg_median_twh, std_twhR, std_twhL, avg_std_twh, MAD_twhR, MAD_twhL, avg_MAD_twh | The thumb-wrist-hand angle is calculated for each frame by computing the arctangent of the ratio betwen the thumb-to-hand length and wrist-to-hand length. It approximates changes in thumb configuration relative to the hand segment. Note that this does not represent a true anatomical joint angle. | 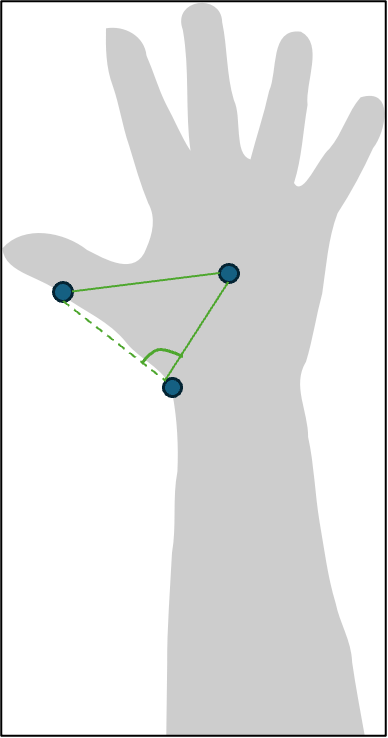 |
| Knee displacement | mean_kneeR, mean_kneeL, avg_mean_knee, median_kneeR, median_kneeL, avg_median_knee, std_kneeR, std_kneeL, avg_std_knee, MAD_kneeR, MAD_kneeL, avg_MAD_knee | Knee displacement was calculated frame-to-frame by computing the Euclidean distance between consecutive (x,y) coordinates of the knee near the patella |  |

Each row defines a group of related behavioral kinematic features considered as inputs for the delirium classification algorithm, how they were calculated, and any references. Variable names follow the convention <summary statistic>_<extremity>_<laterality> for unilateral measures (e.g., right or left), or avg_<summary statistic>_<extremity> for bilateral measures representing the average of the right and left sides. Summary statistics include the mean, median, standard deviation (std), and mean absolute deviation (MAD).

Supplementary Table 5 Model Hyperparameters

| **Model** | **Hyperparameters** |
| --- | --- |
| SVM | - 'class_weight': 'balanced' - 'kernel': 'sigmoid' |
| Logistic Regression | - 'class_weight': 'balanced' - 'penalty': 'elasticnet' - 'solver': 'saga' |
| Gradient Boosting (XGB) | - 'objective' = 'binary:logistic' - 'max_depth': 2 |
| Random Forests | - 'class_weight': 'balanced' - 'max_depth': 3 |

Supplementary Table 6. Keypoint Sensitivity and Specificity by Algorithm, Body Region, and Dataset

|  |  | **Restricted Dataset** | | **Comprehensive Dataset** | |
| --- | --- | --- | --- | --- | --- |
| **Model** | **Region** | **Sensitivity** | **Specificity** | **Sensitivity** | **Specificity** |
| DeepLabCut | Face | 0.90  [0.88, 0.92] | 0.77  [0.71, 0.82] | 0.97  [0.95, 0.99] | 0.74  [0.69, 0.78] |
| FaceMesh | Face | 0.78  [0.69, 0.85] | 0.39  [0.26, 0.54] | 0.74  [0.60, 0.84] | 0.71  [0.61, 0.79] |
| DeepLabCut | Extremity | 0.39  [0.32, 0.46] | 0.96  [0.95, 0.98] | 0.82  [0.78, 0.86] | 0.97  [0.94, 0.98] |
| BlazePose | Extremity | 0.76  [0.70, 0.82] | 0.47  [0.36, 0.59] | 0.78  [0.70, 0.84] | 0.43  [0.28, 0.59] |

Sensitivity and specificity of different keypoint identification models based on the body region (face or extremity) and dataset (restricted or comprehensive). Only non-training images were used to calculate these statistics. Given the clustered nature of the data, a sandwich estimator of the variance was used to calculate 95% confidence intervals, using a logistic regression model with clustering by image (see Methods).

Supplementary Table 7. Keypoint Sensitivity and Specificity of the DeepLabCut Model by Gender and Body Region

| **Region** | **Gender** | **Sensitivity** | **Specificity** |
| --- | --- | --- | --- |
| Extremity | F | 0.796 (293/368) | 0.978 (354/362) |
| Extremity | M | 0.844 (442/524) | 0.956 (302/316) |
| Face | F | 0.990 (1418/1432) | 0.770 (134/174) |
| Face | M | 0.961 (1595/1659) | 0.709 (134/189) |

Sensitivity and specificity of the comprehensive DeepLabCut model based on gender (female or male) and body region (face or extremity).

Supplementary Table 8. Keypoint Sensitivity and Specificity of the DeepLabCut Model by Delirium Status and Body Region

| **Region** | **Delirium Status** | **Sensitivity** | **Specificity** |
| --- | --- | --- | --- |
| Extremity | Negative | 0.830 (621/748) | 0.968 (486/502) |
| Extremity | Positive | 0.792 (114/144) | 0.966 (170/176) |
| Face | Negative | 0.973 (2415/2482) | 0.724 (194/268) |
| Face | Positive | 0.982 (598/609) | 0.779 (74/95) |

Sensitivity and specificity of the comprehensive DeepLabCut model by delirium status (positive or negative) and body region (face or extremity).

Supplementary Table 9. Univariate Behavioral Feature Differences

|  | **Patients with Delirium** | | **Patients without Delirium** | |  |
| --- | --- | --- | --- | --- | --- |
| **Feature Name** | **Median** | **IQR** | **Median** | **IQR** | ***P*** |
| % Eyes Open | 95.11 | 16.79 | 99.68 | 1.93 | 0.029 |
| % Right Eye Open | 93.95 | 29.39 | 98.41 | 9.71 | 0.045 |
| % Left Eye Open | 85.91 | 65.92 | 98.04 | 11.27 | 0.026 |
| Mean Upper Lip Movement | 0.39 | 0.13 | 0.52 | 0.27 | 0.005 |
| Average Mouth Movement | 0.45 | 0.15 | 0.58 | 0.29 | 0.042 |
| Upper Lip Movement Median Absolute Deviation | 0.13 | 0.05 | 0.17 | 0.09 | 0.027 |
| Mean Right Pupil Displacement | 0.36 | 0.15 | 0.47 | 0.29 | 0.020 |
| Mean Left Pupil Displacement | 0.38 | 0.17 | 0.48 | 0.27 | 0.010 |
| Average Mean Pupil Displacement | 0.36 | 0.15 | 0.48 | 0.26 | 0.012 |
| Right Pupil Displacement Median Absolute Deviation | 0.12 | 0.06 | 0.15 | 0.09 | 0.029 |
| Left Pupil Displacement Median Absolute Deviation | 0.12 | 0.07 | 0.16 | 0.09 | 0.010 |
| Average Pupil Displacement Median Absolute Deviation | 0.12 | 0.06 | 0.15 | 0.09 | 0.014 |
| Median Left Horizontal Eyelid Axis | -7.13 | 3.81 | -8.32 | 2.85 | 0.018 |
| Vertical Mouth Axis Standard Deviation | 2.78 | 2.21 | 2.03 | 1.24 | 0.041 |
| Vertical Mouth Axis Median Absolute Deviation | 1.66 | 1.37 | 1.24 | 0.74 | 0.020 |
| Average Mouth Distance Median Absolute Deviation | 1.81 | 1.07 | 1.35 | 0.69 | 0.016 |
| Left Blink Rate Standard Deviation | 18.08 | 76.99 | 4.94 | 12.97 | 0.23 |
| Head Pitch Standard Deviation | 0.37 | 0.14 | 0.47 | 0.28 | 0.009 |
| Head Yaw Standard Deviation | 0.42 | 0.15 | 0.59 | 0.40 | 0.002 |
| Mean Right Wrist Displacement | 2.28 | 25.04 | 1.04 | 2.35 | 0.032 |
| Median Chest Openness Area | 0.60 | 0.12 | 0.54 | 0.07 | 0.013 |
| Chest Openness Area Median Absolute Deviation | 0.01 | 0.01 | 0.00 | 0.01 | 0.014 |
| Median Left Wrist-Shoulder-Elbow Angle | 0.17 | 0.72 | 0.62 | 0.73 | 0.044 |

Exploratory univariate comparisons of video-based behavioral features significantly different between patients with or without delirium are shown in the table. Statistical significance was defined to be a p-value below 0.05 and was calculated using the Mann Whitney U-Test, which compared the median feature values between patients with or without delirium. P-values were not corrected for multiple comparisons given the exploratory and descriptive nature of this presentation.

Supplementary Table 10. Results of Different Classification Model and Feature Selection Combinations

Algorithms include LR = Logistic Regression; RF = Random Forests; SVM = Support Vector Machine; XGB = XGBoost. Feature Selection methods include None: all features included, Chi square test, Lasso Regression, mRMR: minimal Redundancy, Maximal Relevance, and MI = Mutual Information.

Additional Abbreviations: ROC AUC = Receiver Operating Characteristic Area Under the Curve. Std = Standard Deviation. Sens = Sensitivity. Spec = Specificity. The row highlighted in green (SVM, mRMR, 40), represents the best performing model.

| **Algorithm** | **Feature Selection** | **Number of Features** | **Mean ROC AUC** | **Std. ROC AUC** | **Mean Sens.** | **Std. Sens.** | **Mean Spec.** | **Std. Spec.** |
| --- | --- | --- | --- | --- | --- | --- | --- | --- |
| LR | None | 232 | 0.71 | 0.13 | 0.48 | 0.22 | 0.86 | 0.06 |
| LR | Chi^2^ | 10 | 0.66 | 0.15 | 0.82 | 0.14 | 0.25 | 0.14 |
| LR | Chi^2^ | 20 | 0.66 | 0.16 | 0.78 | 0.20 | 0.39 | 0.15 |
| LR | Chi^2^ | 40 | 0.65 | 0.15 | 0.61 | 0.24 | 0.62 | 0.19 |
| LR | Chi^2^ | 80 | 0.68 | 0.12 | 0.51 | 0.20 | 0.82 | 0.08 |
| LR | Lasso | Variable | 0.71 | 0.12 | 0.85 | 0.15 | 0.26 | 0.13 |
| LR | mRMR | 10 | 0.73 | 0.12 | 0.67 | 0.24 | 0.65 | 0.22 |
| LR | mRMR | 20 | 0.74 | 0.11 | 0.60 | 0.18 | 0.81 | 0.09 |
| LR | mRMR | 40 | 0.76 | 0.10 | 0.60 | 0.20 | 0.84 | 0.06 |
| LR | mRMR | 80 | 0.72 | 0.12 | 0.55 | 0.20 | 0.84 | 0.06 |
| LR | MI | 10 | 0.56 | 0.13 | 0.37 | 0.24 | 0.70 | 0.24 |
| LR | MI | 20 | 0.55 | 0.13 | 0.36 | 0.22 | 0.76 | 0.18 |
| LR | MI | 40 | 0.59 | 0.13 | 0.38 | 0.20 | 0.80 | 0.08 |
| LR | MI | 80 | 0.64 | 0.13 | 0.45 | 0.20 | 0.83 | 0.08 |
| RF | None | 232 | 0.71 | 0.09 | 0.04 | 0.08 | 0.99 | 0.02 |
| RF | Chi^2^ | 10 | 0.64 | 0.15 | 0.14 | 0.14 | 0.95 | 0.04 |
| RF | Chi^2^ | 20 | 0.64 | 0.14 | 0.09 | 0.10 | 0.96 | 0.04 |
| RF | Chi^2^ | 40 | 0.69 | 0.09 | 0.08 | 0.09 | 0.97 | 0.03 |
| RF | Chi^2^ | 80 | 0.70 | 0.11 | 0.06 | 0.08 | 0.98 | 0.03 |
| RF | Lasso | Variable | 0.72 | 0.11 | 0.09 | 0.10 | 0.96 | 0.04 |
| RF | mRMR | 10 | 0.68 | 0.12 | 0.11 | 0.10 | 0.96 | 0.04 |
| RF | mRMR | 20 | 0.68 | 0.10 | 0.10 | 0.12 | 0.97 | 0.03 |
| RF | mRMR | 40 | 0.68 | 0.10 | 0.08 | 0.09 | 0.97 | 0.03 |
| RF | mRMR | 80 | 0.67 | 0.11 | 0.07 | 0.10 | 0.99 | 0.02 |
| RF | MI | 10 | 0.56 | 0.11 | 0.05 | 0.08 | 0.95 | 0.06 |
| RF | MI | 20 | 0.58 | 0.11 | 0.03 | 0.06 | 0.96 | 0.05 |
| RF | MI | 40 | 0.61 | 0.11 | 0.04 | 0.06 | 0.97 | 0.04 |
| RF | MI | 80 | 0.64 | 0.10 | 0.02 | 0.05 | 0.98 | 0.03 |
| SVM | None | 232 | 0.79 | 0.09 | 0.71 | 0.16 | 0.78 | 0.07 |
| SVM | Chi^2^ | 10 | 0.50 | 0.18 | 0.55 | 0.22 | 0.60 | 0.12 |
| SVM | Chi^2^ | 20 | 0.57 | 0.19 | 0.65 | 0.20 | 0.62 | 0.11 |
| SVM | Chi^2^ | 40 | 0.62 | 0.16 | 0.65 | 0.18 | 0.63 | 0.12 |
| SVM | Chi^2^ | 80 | 0.71 | 0.10 | 0.70 | 0.17 | 0.67 | 0.11 |
| SVM | Lasso | Variable | 0.61 | 0.14 | 0.62 | 0.21 | 0.63 | 0.11 |
| SVM | mRMR | 10 | 0.52 | 0.17 | 0.52 | 0.23 | 0.59 | 0.10 |
| SVM | mRMR | 20 | 0.62 | 0.16 | 0.63 | 0.23 | 0.65 | 0.11 |
| SVM | mRMR | 40 | 0.69 | 0.13 | 0.69 | 0.18 | 0.66 | 0.10 |
| SVM | mRMR | 80 | 0.73 | 0.09 | 0.76 | 0.14 | 0.67 | 0.11 |
| SVM | MI | 10 | 0.47 | 0.14 | 0.39 | 0.26 | 0.63 | 0.27 |
| SVM | MI | 20 | 0.53 | 0.15 | 0.41 | 0.24 | 0.66 | 0.20 |
| SVM | MI | 40 | 0.57 | 0.15 | 0.50 | 0.25 | 0.69 | 0.12 |
| SVM | MI | 80 | 0.68 | 0.12 | 0.61 | 0.20 | 0.74 | 0.10 |
| XGB | None | 232 | 0.67 | 0.11 | 0.12 | 0.14 | 0.95 | 0.05 |
| XGB | Chi^2^ | 10 | 0.64 | 0.14 | 0.16 | 0.14 | 0.92 | 0.06 |
| XGB | Chi^2^ | 20 | 0.62 | 0.12 | 0.13 | 0.13 | 0.92 | 0.06 |
| XGB | Chi^2^ | 40 | 0.61 | 0.10 | 0.15 | 0.13 | 0.92 | 0.05 |
| XGB | Chi^2^ | 80 | 0.62 | 0.12 | 0.16 | 0.12 | 0.92 | 0.07 |
| XGB | Lasso | Variable | 0.69 | 0.09 | 0.18 | 0.15 | 0.91 | 0.06 |
| XGB | mRMR | 10 | 0.63 | 0.12 | 0.18 | 0.14 | 0.91 | 0.07 |
| XGB | mRMR | 20 | 0.61 | 0.11 | 0.19 | 0.14 | 0.92 | 0.06 |
| XGB | mRMR | 40 | 0.61 | 0.13 | 0.17 | 0.13 | 0.91 | 0.06 |
| XGB | mRMR | 80 | 0.60 | 0.12 | 0.16 | 0.13 | 0.93 | 0.06 |
| XGB | MI | 10 | 0.59 | 0.13 | 0.15 | 0.14 | 0.90 | 0.07 |
| XGB | MI | 20 | 0.61 | 0.13 | 0.12 | 0.12 | 0.92 | 0.06 |
| XGB | MI | 40 | 0.64 | 0.12 | 0.15 | 0.14 | 0.93 | 0.06 |
| XGB | MI | 80 | 0.65 | 0.10 | 0.13 | 0.12 | 0.94 | 0.05 |
